# Evaluation of Oxford Nanopore Technologies workflows for genomic epidemiology of outbreak‐associated bacterial isolates in the clinical setting

**DOI:** 10.1101/2025.04.25.25326448

**Authors:** Stefan Neuenschwander, Loïc Borcard, Sonja Gempeler, Miguel Terrazos Miani, Carlo Casanova, Alban Ramette

**Affiliations:** Institute for Infectious Diseases, University of Bern, Bern, Switzerland; Multidisciplinary Center for Infectious Diseases, University of Bern, Bern, Switzerland

**Keywords:** Oxford Nanopore Technologies, R10.4.1, genomic Epidemiology, outbreak, bacterial pathogens, clinical setting, cgMLST, plasmids, genome assembly

## Abstract

Accurate and efficient whole-genome sequencing (WGS) is crucial for clinical diagnostics and surveillance of bacterial infections. Here, we investigate the potential of a new Oxford Nanopore Technologies (ONT) workflow for WGS of clinically relevant bacterial isolates. Specifically, we assess the performance of R10.4.1 flow cells in combination with the V14 version of the transposase-based (RBK) library preparation kit to provide rapid and accurate genomic epidemiological comparisons of bacterial species of clinical importance. We focused on retrospective collections of outbreak-associated *Corynebacterium diphtheriae* (CDIP) and vancomycin-resistant Enterococci (VRE), and benchmarked expected performance parameters such as genome assembly quality, genotyping (MLST, cgMLST), SNP profiling and antimicrobial resistance and virulence prediction, against WGS data obtained routinely by Illumina MiSeq sequencing. Complete concordance with Illumina results was observed for MLST in both species, and for cgMLST in CDIP, across all ONT kits and software evaluated. For VRE, however, cgMLST results varied with strain identity, library preparation kit, and analysis parameters, likely due to software challenges to correctly call methylated bases. Yet, the use of the latest basecalling models combined with PCR-based library preparation kit (RPB) reliably reproduced Illumina cgMLST results across all tested VRE strains. By testing two hybrid strategies combining PCR-free and PCR-based library preparation approaches, we also showed that combining PCR-free and PCR-based methods may yield a promising strategy, achieving both high accuracy and assembly completeness. Genomic-based AMR prediction was consistent across sequencing methods, and we further highlight advantages and limitations of the PCR-based, PCR-free, and mixed assemblies, to inform on the genomic context of AMR genes. This study demonstrates that a Nanopore-only sequencing approach may offer improved accuracy and consistency for classical bacterial typing in outbreak investigations, paving the way to wider use in clinical microbiology laboratories.

## 1 Introduction

Whole-genome sequencing (WGS) has become the gold standard for bacterial strain typing, revolutionizing our ability to elucidate infection-epidemiological connections due to its high discriminatory power (Van Goethem et al., 2019). Consequently, WGS is increasingly employed in clinical settings to enhance outbreak detection, implement epidemiological surveillance, and improve infection control, leveraging its unparalleled resolution (Didelot et al., 2012; Price et al., 2023; Schadron et al., 2024; Simar et al., 2021). Common approaches for assessing genetic relatedness from WGS data include gene-by-gene comparisons, such as core-genome multi-locus sequence typing (cgMLST) or whole-genome multi-locus sequence typing (wgMLST), and single-nucleotide polymorphism (SNP) analysis (Deurenberg et al., 2017). While SNP typing offers high resolution, it is computationally intensive and time-consuming (Higgs et al., 2022; Nadon et al., 2017). In contrast, cgMLST-typing provides a standardized nomenclature with lower computational demands (Deurenberg et al., 2017). Target-free, k-mer based methods, like split k-mer analysis (SKA), are emerging but are not yet standard for transmission analysis (Harris, 2018; Higgs et al., 2022). Despite the widespread use of these methods, a standardized approach for interpreting WGS data in epidemiological investigations remains undefined (Higgs et al., 2022).

Healthcare facilities have traditionally relied on short-read Illumina technology for WGS and genomic pathogen surveillance (Greig et al., 2019; Schürch et al., 2018). However, long-read next-generation sequencing (NGS), particularly provided by Oxford Nanopore Technologies (ONT)(Deamer et al., 2016), is gaining traction. ONT sequencing offers rapid turnaround times, real-time data, long-read capabilities, and lower capital costs, potentially reducing outbreak investigation times or enabling genomic surveillance in resource-limited settings (Sanderson et al., 2023; Wagner et al., 2023). Studies have shown promising results when using ONT sequencing for bacterial sequence typing and epidemiology (Ahrenfeldt et al., 2017; Magi et al., 2018), including largely consistent findings with Illumina for vancomycin-resistant (Oh et al., 2022), *Mycobacterium tuberculosis* (Hall et al., 2023), *Staphylococcus aureus* (Liao et al., 2022) and methicillin-resistant *Staphylococcus aureus* (MRSA) (Ferreira et al., 2021). Several studies also demonstrate successful Nanopore-based typing in *Salmonella* (Wu et al., 2022; Xian et al., 2022), with one addressing homopolymer error reduction and combined Illumina/Nanopore analysis (Xian et al., 2022). Smaller studies also report consistent results between Illumina and Nanopore for highly pathogenic bacteria (Linde et al., 2023), *E. coli* (Greig et al., 2019), VRE (Tarumoto et al., 2017), improved resolution of hospital VRE isolates (Both et al., 2022) and *Klebsiella pneumoniae* strain typing (Cao et al., 2016).

Despite significant improvements in ONT technology, including higher throughput and lower error rates, and increasing use for bacterial genome assembly (Dilthey et al., 2020; Sereika et al., 2022), challenges remain. These include the sensitivity of established typing methods like multi-locus sequence typing (MLST)(Maiden et al., 1998, 2013), cgMLST, or cgSNP (Schürch et al., 2018) to Nanopore sequencing errors, and the need to further compare new Nanopore data and Illumina-based data for outbreak detection. For clinical diagnostics, where accuracy is key for correct treatment and control measures, these errors can be critical. This re-evaluation is particularly true given the recent improvement in ONT sequencing accuracy due to the development of newer flow cells and basecalling algorithms, making it increasingly reliable for clinical applications (Delahaye and Nicolas, 2021; Linde et al., 2023). For instance, the R10.4.1 flow cell version has improved modal sequencing accuracy of 99.6% over previous flow cell versions, which may be crucial for high-resolution genotyping necessary in clinical diagnostics. In addition, ONT transposase-based library preparation kits (e.g. RBK) now offer very fast turnaround time from isolates to sequencing results. In addition, the possibility to combined PCR and PCR-free library preparation has not been fully assessed in terms of turnaround time, cost, and effects on the genomic sequencing accuracy.

To assess the suitability of the latest ONT V14 RBK kits for genomic epidemiological comparisons, particularly in outbreak-prone, clinically relevant species, we conducted a retrospective, comparative study using clinical isolates of *Corynebacterium diphtheriae* (CDIP) and vancomycin-resistant Enterococci (VRE). Recognizing Illumina short-read sequencing as the current gold standard, renowned for its accuracy but limited in resolving genomic context and structure, we aimed to evaluate whether the ONT long-read platform provide actionable data for genomic epidemiology decision-making, and focused on typing (MLST, cgMLST), resistance (AMR) and plasmid analyses, thereby determined the performance of ONT using two examples of bacterial outbreak investigations.

## 2 Methods

### Sample origin, cultivation, DNA extraction

All CDIP isolates were associated with local outbreaks in Switzerland from July to September 2022, and were described in a previous study (Kofler et al., 2022). The corresponding Illumina MiSeq data is available as BioProject PRJNA889706). VRE isolates, associated with local hospital outbreaks in a Swiss hospital, were collected between 2017 and 2021. They were isolated on CHROMagar VRE plates (CHROMagar, Paris, France) during routine analysis by the clinical microbiology laboratory of the Institute for Infectious Diseases (IFIK), University of Bern. Species identification was determined by matrix-assisted laser desorption ionization–time of flight (Bruker Daltonics, Bremen, Germany). Susceptibility testing for all isolates was performed during routine clinical processing at IFIK. All isolates were stored at -80°C, and re-grown on CSBA plates before extraction. Genomic DNA was extracted from agarose plates using PureLink Genomic DNA kit (Thermofisher, Switzerland), or with Maxwell RSC Cultured Cells DNA Kit (Promega, Switzerland).

### Whole Genome Sequencing

NGS libraries for Illumina sequencing were produced with the Nextera DNA Flex Library Prep kit (Illumina, Switzerland) according to the manufacturer’s recommendations, and sequenced on an Illumina MiSeq sequencer with v2 reagents in 2x 150 paired-end mode, at the Next Generation Sequencing Platform of the Institute for Infectious Diseases, Bern. Nanopore sequencing libraries were produced with the rapid barcoding chemistry SQK-RBK114.96 (RBK), and Rapid PCR barcoding chemistry SQK-RPB114.96 (RPB) according to the manufacture’s recommendations. The libraries were loaded onto standard GridION flowcells (FLO-MIN114) and sequenced in batches of 21 samples for 12-72 hours on GridION X5 sequencers with real-time basecalling and under high accuracy mode (HAC). The raw signal files (fast5_pass/pod5_pass directories) were subsequently re-basecalled either on the GridION, or with standalone basecallers on a separate Linux workstation. All software versions and basecaller models are provided in the supplementary material (Table S1).

### Ethics statement

All CDIP and VRE isolates in the present study have been anonymized and no patient information is used in the interpretation of the bacterial genomic data. Publication of this analysis does not harm or influence neither patients nor institutions. Ethical committee approval was therefore not requested.

### Bioinformatic analyses

#### Genome assembly and polishing

All genomes were reconstructed with the same software versions and parameters, except for the assembly parameters and for the models used for genome polishing, which were adjusted to the basecalling mode (SUP or HAC), and the basecalling model respectively (**Figure 1**). All reads were filtered with *Nanofilt* (version 2.8.0, parameters: -q 10 -l 500 --headcrop 30 --tailcrop 30) (De Coster et al., 2018), assembled with *Flye* (version 2.9.2) (Kolmogorov et al., 2019), and polished with *Medaka* (version 1.11.3) (“nanoporetech/medaka” 2024). Detailed information on basecalling parameters, models and assemblies can be found in the supplementary material (**Table S1**). To mask bases with ambiguous base compositions in the corresponding reads, selected assemblies were processed with the software MPOA (Lohde et al., 2024).

**Figure 1.**
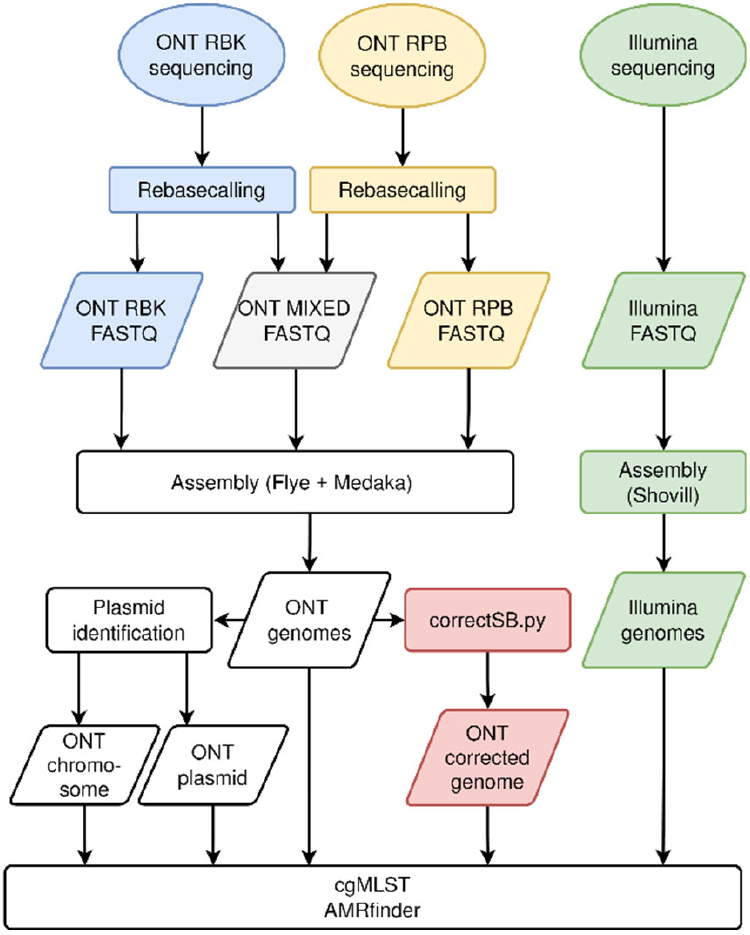
Overview of the bioinformatic analyses.

#### Combining PCR-based and PCR-free Nanopore reads

To combine superior assembly performance of RBK with the robustness against single base errors caused by methylation of RPB, two different approaches were tested: a) **Read mixtures**. Random subsets of the reads produced with RBK and RPB were combined at theoretical coverage depths of 70x and 30X respectively, to obtain mixed datasets with total coverage depths of 100X. These read sets were processed as described above for the non-mixed datasets. b) **Assembly correction**. Reads sets produced with RBK and RPB were mapped separately against an assembly produced from the RBK reads with the software *minimap2* (version. 2.24) (Li, 2018). The resulting SAM files were sorted (*samtools*, version 1.15.1) (Danecek et al., 2021) and further analyzed with the software *Pysamstats* (version 1.1.2) (Miles, 2024) to obtain per-strand base compositions for each assembly position.

These base compositions were then processed using a custom Python script as follows (**Figure 2**): 1) We assigned a confidence level (high or low) to each base of the original RBK assembly, based on the underlying base compositions of the mapped reads. Assembly positions were considered as “low confidence” if one or more of the following criteria applied: Low read coverage (<20X coverage depth), mixed base composition (<80% of the reads matching the most frequent base), or strand bias (different dominant bases between the forward and reverse strand). Otherwise, the positions were considered as “high confidence”. 2a) The RPB majority consensus base was obtained for each base position of the original assembly. 2b) We assigned a confidence level (high or low) to the RPB majority consensus bases, using the same criteria described in step 1 above. 3) The assembly was corrected for low confidence positions in the original assembly, which were replaced by the majority consensus of the RPB reads if the confidence was high for the latter, and by N, if the confidence in the RPB read set was low too. Each “high confidence position” of the original assembly was kept.

**Figure 2.**
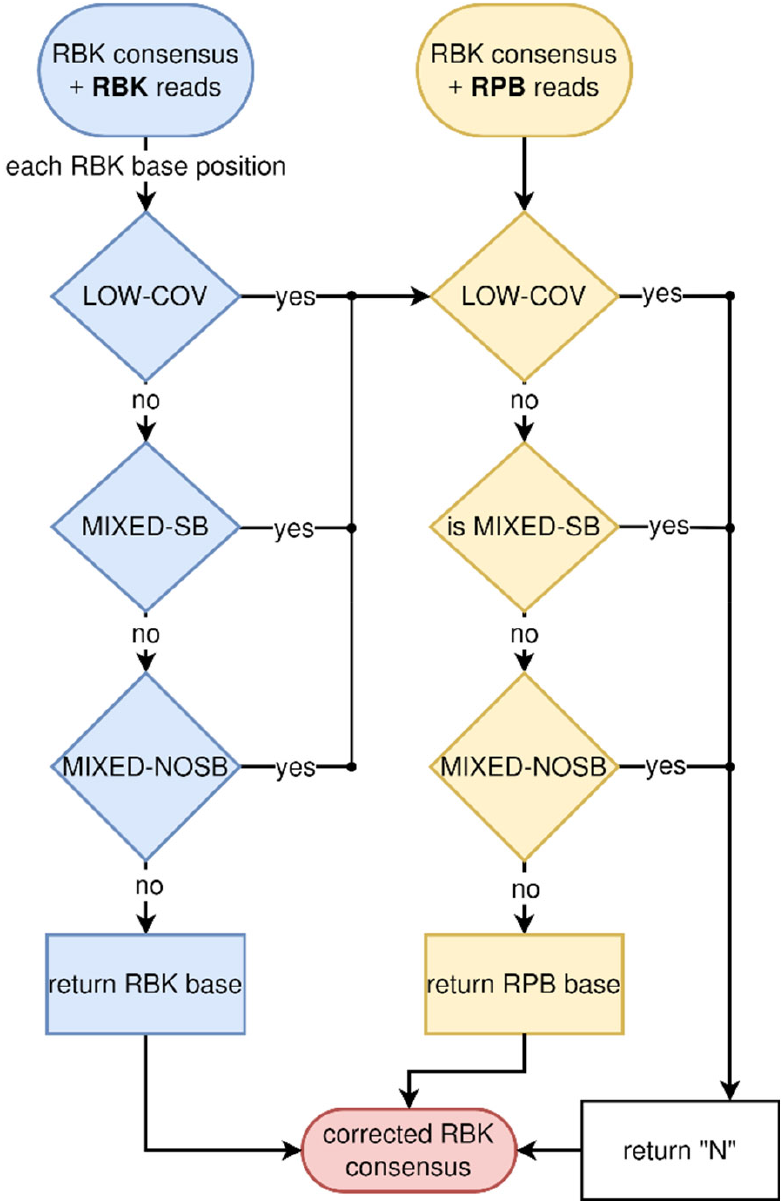
Correction algorithm of RBK-based assembly using RPB reads. Abbreviations: LOW-COV, coverage below the threshold of 20X; MIXED-SB, mixed base composition where the majority base constitutes more than 66% of all bases on one strand, and less than 33% on the other strand; MIXED-NOSB, mixed bases that do not fulfil the criteria for strand bias mentioned above.

#### Typing and comparison of the different treatments

All assemblies were imported into *SeqSphere*+ (v. 8.4.0; Ridom GmbH, Germany) and processed with the cgMLST schemes *E. faecium* cgMLST v1.1 (de Been et al., 2015), and a custom task template encompassing 1,319 core genes for CDIP based on the Institute Pasteur scheme, as described in (Kofler et al., 2022). Comparison tables (containing allele ids for each target gene and assembly) were created for the Nanopore and Illumina assemblies of each analyzed species and exported for further processing with custom python and R scripts.

#### AMR and plasmid analyses

Annotations and predicted proteins were generated with the software *Prokka* (version 1.14.6) (Seemann, 2014) and used as input for *AMRFinderPlus* (version 3.11.2, database: 2023-08-08.2, parameters -p -g -n --annotation_format prokka --organism) (Feldgarden et al., 2021). For plasmid analysis, WGS reads from treatments ‘SUPD43’, ‘SUPDP43’, and ‘SUPD&P43’ were reassembled and analyzed with the “long read” module with default parameters (*SeqSphere*+; version 10.0.0). Assembly statistics (number of replicons, circularity) were extracted from the result section “Chromosome & Plasmid Overview”, resistances and plasmid clusters (single linkage, max mash distance: 0.001, distance added per 1% size difference 0.0003, max allowed size difference: 40%) were based on a local database created via “Create Task Template for Plasmid Mash Database” in *SeqSphere*+. The resulting output tables were analyzed using Python custom scripts.

#### Data and code availability

Genome assemblies, Illumina MiSeq and ONT data are available from BioProject PRJNA889706 (http://www.ncbi.nlm.nih.gov/bioproject/889706), and BioProject PRJNA1230056 (http://www.ncbi.nlm.nih.gov/bioproject/1230056). The correction script (Figure 2) is available at: https://github.com/RametteLab/NanoporeHybridKP

## 3 Results

### VRE genomic epidemiology using Nanopore vs. Illumina WGS

We choose a representative and diverse collection of clinical VRE isolates, consisting of six different MLST sequence types, three clonal clusters, and 12 different cgMLST complex types based on Illumina assemblies (**Figure 3A**). In terms of target genes recovery, we observed near identical performance between Illumina and the top-performing Nanopore WGS methods: Percentages of successfully detected cgMLST target genes in the assemblies were highest for Illumina (98.80%), closely followed by the PCR-based Nanopore approach SUPDP43 (98.77%) and the most recent PCR-free approach SUPD50 (98.71%). The lowest percentage was found for the HAC43.masked (97.46%), which was still well above the quality threshold of 90% (**Figure 3B**).

**Figure 3.**
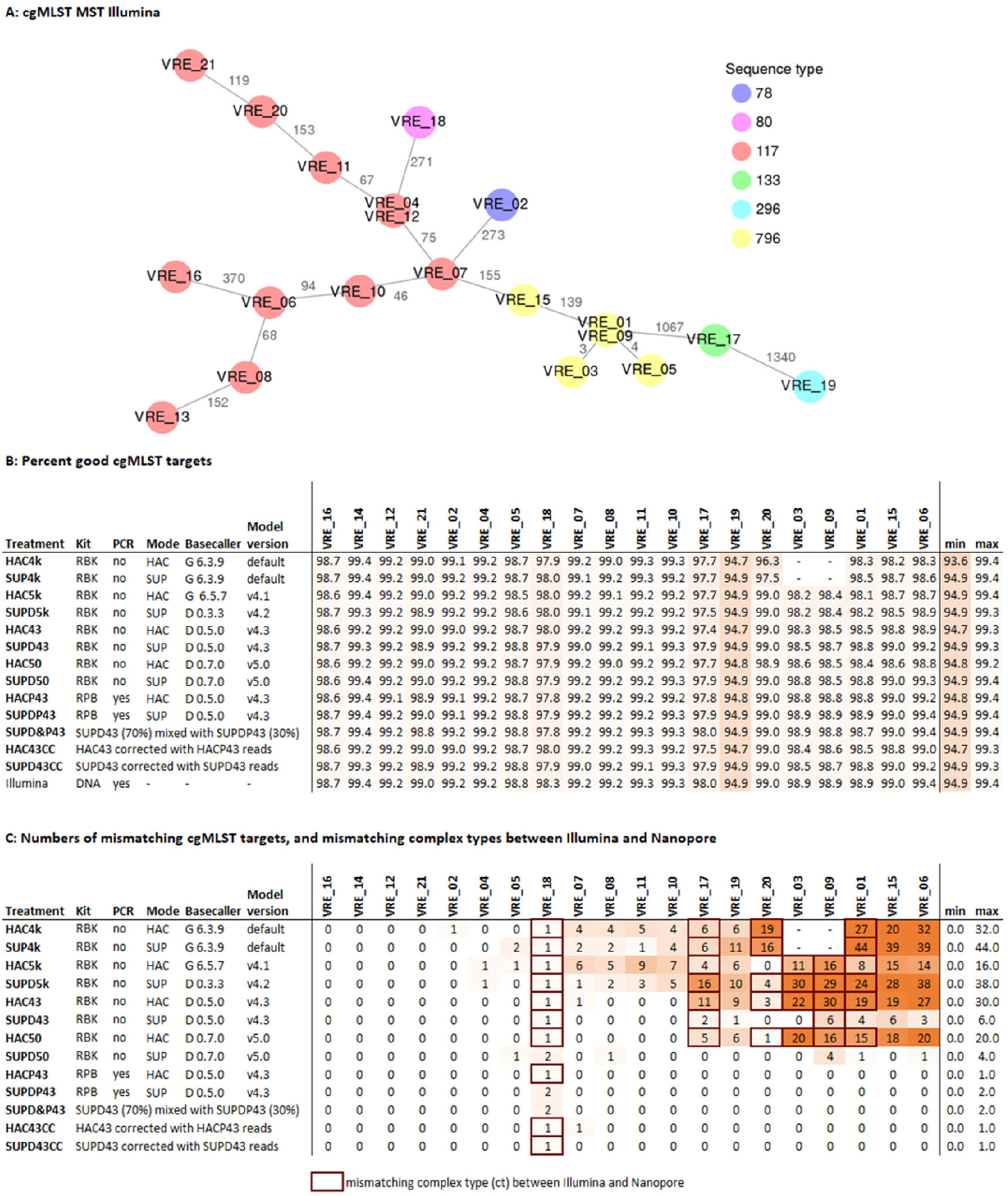
Comparison of cgMLST results for clinical VRE isolates sequenced with Illumina or Nanopore (ONT). The cgMLST scheme contained 1,423 target genes. A) Minimum spanning tree of the 20 Illumina assemblies. B) Percentage the 1,423 cgMLST target genes that were detected (and passed the QC) in the different assemblies. C) Numbers of mismatching alleles between the ONT and corresponding Illumina assemblies, and mismatching complex types between the two technologies (indicated by red boxes). Letters G and D in the column Basecaller stand for Guppy and Dorado, respectively. Detailed explanation about treatments is provided in **Table S1**.

A near-perfect agreement was found between Illumina and the top-performing Nanopore methods for the individual cgMLST allele assignments. The numbers of mismatching alleles between the Illumina and Nanopore assemblies varied strongly between isolates, Nanopore software versions, basecalling models, as well as the library preparation kits: For VRE21, for example, no mismatching alleles were detected, irrespective of the approach used to create the Nanopore assemblies, whereas 0 to 44 mismatching alleles were detected in the assemblies produced from isolate VRE01 (**Figure 3C**). Noticeably, PCR-based library preparation outperformed PCR-free alternatives irrespective of the basecalling models used, allowing for a perfect replication of the allelic profiles obtained with Illumina in all but one case, with a maximum number of two mismatches (VRE18). Mixtures of PCR-based and PCR-free reads, as well as PCR-free assemblies that were corrected with PCR-based reads performed on par PCR-free reads in these aspects.

The PCR-free approach was also more sensitive to the analysis parameters: SUP models clearly outperformed HAC models, and the most recent iterations of the software (and models) outperformed previous versions. The best combination of parameters resulted in a perfect replication of the allelic profiles in 16 out of 20 cases with a maximum of four mismatches (**Figure 3C**). Subsequent masking with the software *MPOA* resulted in the exclusion of all but one of the previously mentioned mismatching alleles (*SeqSphere*+ excludes genes with N’s from the analysis), while the percentage of successfully detected cgMLST targets dropped by 0.8%, remaining well above the threshold of 90% (SUPD34: 98.68%, SUP43.masked: 97.90%; **Figures S1A, S1B**).

MLST classification into Sequence Type (ST) was successful across all tested combinations that did not involve masking with MPOA (**Figure S1D**). The latter resulted in two unassigned STs in the treatment HAC43.masked (**Figure S1D**), whereas treatment SUP43.masked did not result in a wrong ST assignment. cgMLST classification into complex types (CTs) was more sensitive to the variability introduced by the different treatments: The PCR-based treatments SUPDP43 and HACP43 matched the Illumina CT’s in all, and in 20 out of 21 cases respectively. The single CT 1552 (instead of CT 2887) that differed between the PCR-based treatments was a result of one target gene (EFAU004_01770, hypothetical protein), which was exclusively found in SUPDP43 in otherwise identical allelic profiles (**Figure S1D**). The mixtures of PCR-based and PCR-free reads matched the Illumina CT’s in all cases, and the PCR-free assemblies that were corrected with PCR-based reads in 20 out of 21 cases respectively. The single mismatch was caused by the absence of the same target gene, as described above for the PCR-based approaches. Among the PCR-free treatments, SUPD50 performed the best with all 21 correct CT assigned, followed by SUPD43 with 19/21 correct assignments (**Figure 3C**). Subsequent MPOA masking (SUP43.masked) corrected two of the remaining mis-assigned CTs, but another CT was assigned for VRE10, while no CT was assigned for it based on the Illumina assembly (**Figure S1**).

Noticeably, a large proportion of the mismatching bases between Illumina and Nanopore assemblies were found in assembly positions with conflicting information between the forward and reverse reads (**Figure 4A**). This phenomenon is referred to as strand bias (SB) in subsequent sections. We analyzed the frequency of SB as a function of its genomic context and MLST types (**Figure 4B**): In our dataset, SB was more often observed on plasmids than on chromosomes. No distinct pattern related to sequence type (ST) was identified; instead, SB levels appeared to be strain-specific rather than ST-specific (**Figure 4B**).

**Figure 4.**
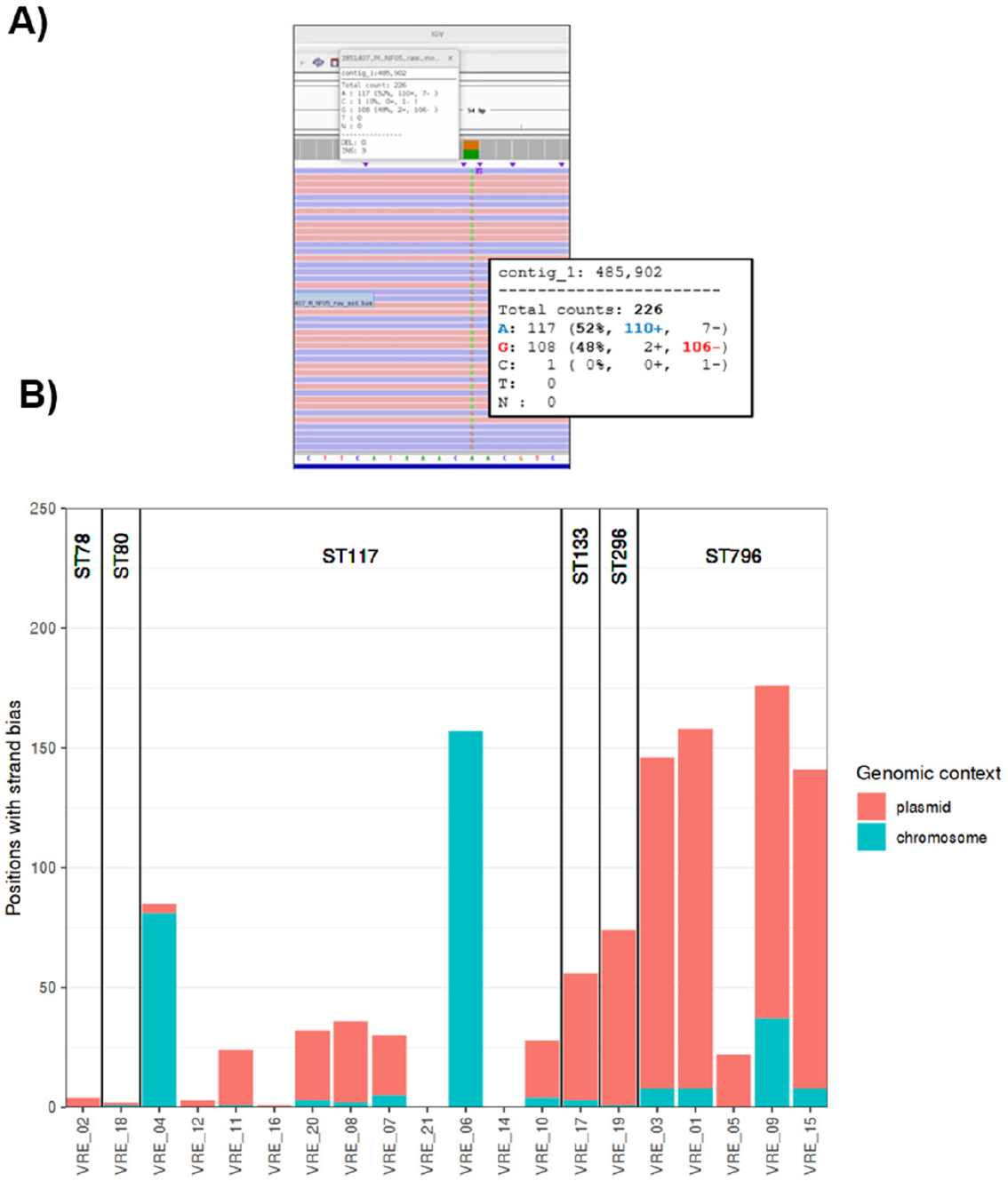
A) Example of an ambiguous base position: The majority of forward and reverse reads differ at the called base. B) Numbers of ambiguous base positions detected by genomic locations and by MLST ST.

### VRE-predicted AMR and plasmids

To evaluate whether ONT assemblies are suitable for AMR analysis, we applied AMRFinderPlus to the Illumina and ONT assemblies: On the class level, predictions for the Illumina and ONT based assemblies matched in all cases (**Figure 5A**). On the antibiotic subclass level, discrepancies were found for one isolate (VRE_02), for which gentamicin and tobramycin resistances were exclusively predicted for the ONT assemblies (**Figure 5B**). On the gene level, the bifunctional aminoglycoside-modifying enzyme *AAC(6′)-Ie– APH(2′′)-Ia*, which confers resistance to all commercially available aminoglycosides except streptomycin (Arias and Murray, 2012), was exclusively detected in the ONT assemblies in five cases (**Figure 5C**). However, in all but one isolate (VRE_02), the C-terminal domain of *AAC(6′)-Ie–APH(2′′)-Ia*, was detected in the Illumina and ONT assemblies, explaining the matching resistance predictions (**Figures 6A, 6B**).

**Figure 5.**
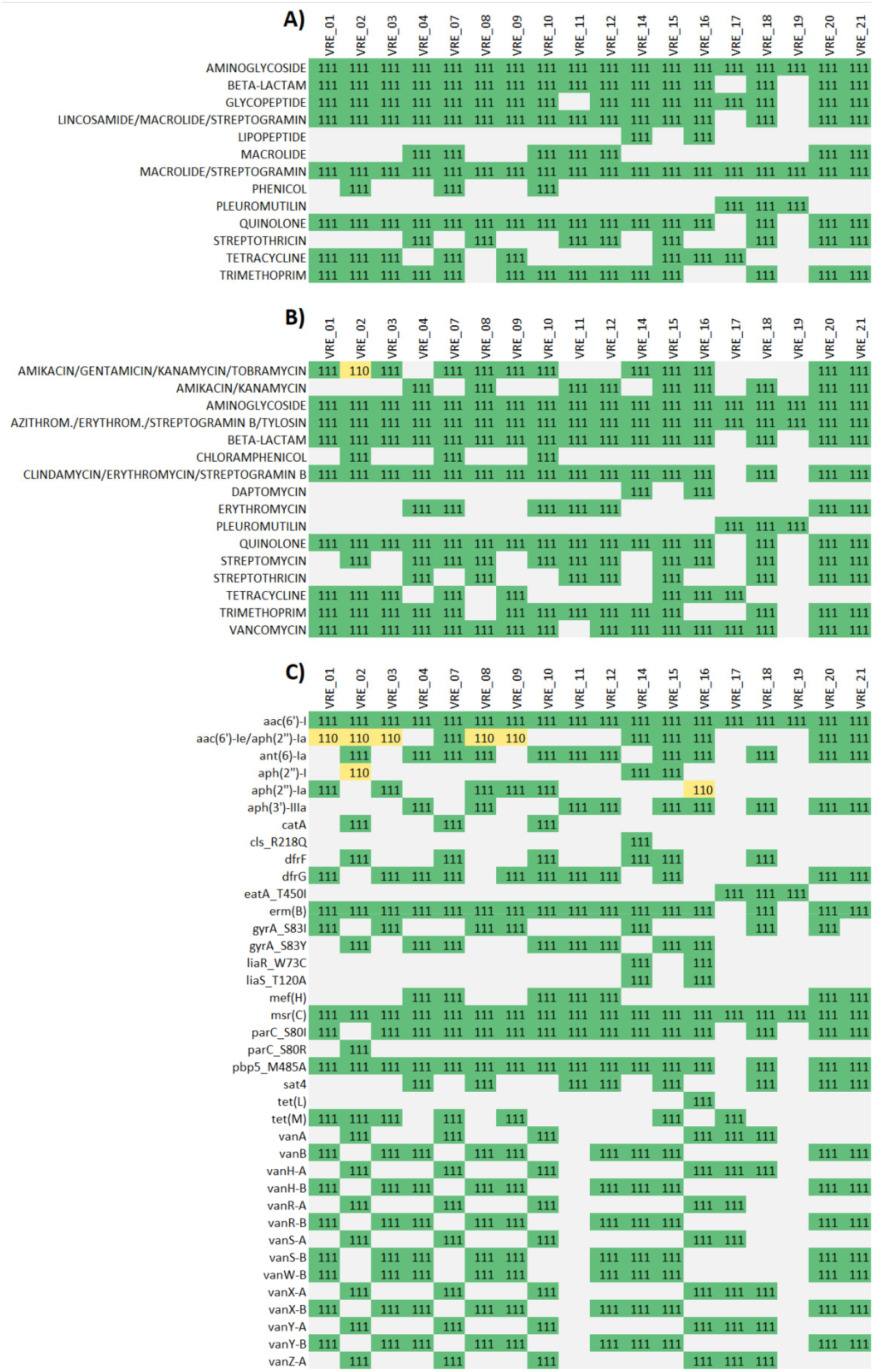
Comparison of the predicted resistance genes based on ONT and Illumina assemblies. Tuples indicate presence (1) or absence (0) in the treatments SUBDP_43 (RPB), SUPD_43 (RBK), and ILLUM (Illumina) with A) antibiotic classes, B) antibiotic subclasses, C) resistance genes.

**Figure 6.**
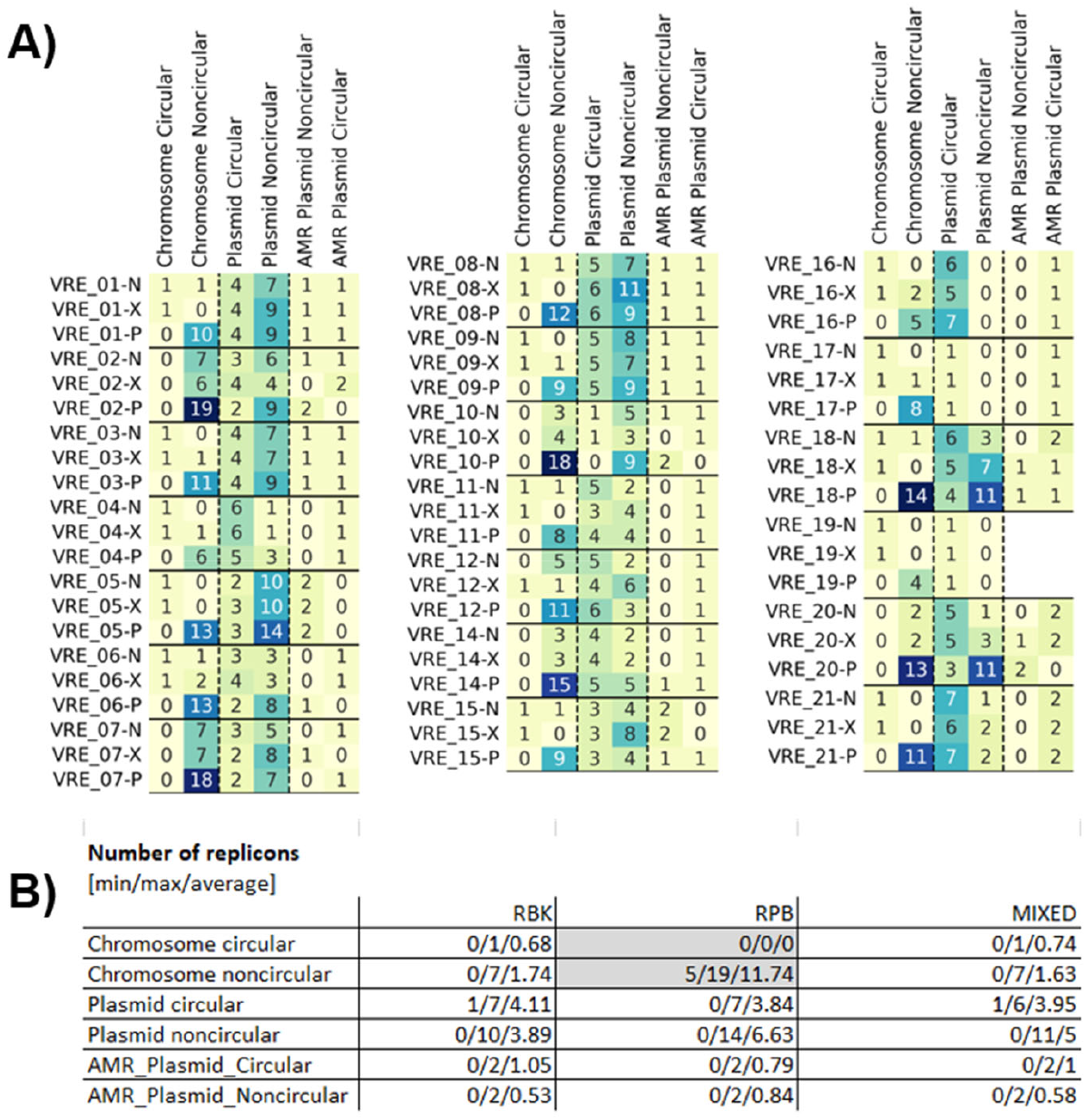
Assembly performance of PCR-based, PCR-free, and mixed Nanopore reads. A) Numbers of contigs by categories based on replicon type, circularity and presence of AMR genes. B) Summary of the data provided in A). Grey shadings indicate significant differences to the other treatments within one category (Mann-Whitney U test; corrected p-value for multiple testing, p<0.05).

Different Nanopore approaches were compared with respect to their performance for plasmid analysis (PCR-free, PCR-based and mixtures of both). PCR-free and mixed Nanopore methods showed superior chromosome assembly performance than PCR-based Nanopore assemblies, as PCR-based Nanopore assemblies showed a higher degree of chromosome fragmentation than PCR-free, or mixed assemblies. Plasmids assembly performance did not differ significantly between the tested methods (**Figure 6**).

Discrepancies in the predicted genomic context of resistance genes were identified in four out of the 19 isolates when comparing the tested Nanopore approaches: Tetracycline resistance in VRE02, along with gentamicin and tobramycin resistance in VRE16, was determined to be plasmid-mediated in PCR-free and mixed assemblies, but not in PCR-based assemblies. Conversely, amikacin, gentamicin, kanamycin, and tobramycin resistance in VRE14 were identified as plasmid-mediated only in the PCR-based assemblies. Vancomycin resistance was predicted to be plasmid-mediated in PCR-based and mixed assemblies of VRE20, but not in the PCR-free variant (**Figure 7**).

**Figure 7.**
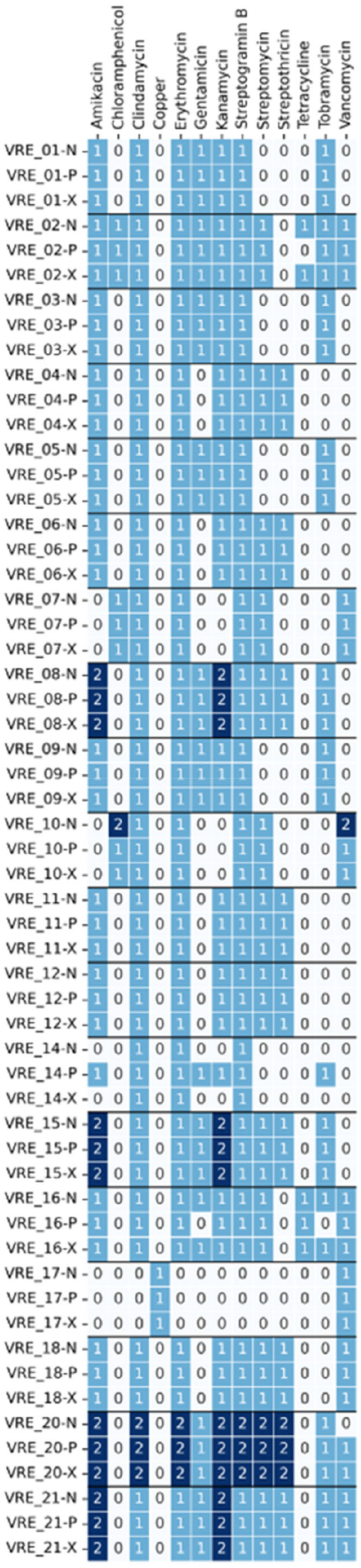
Plasmid-borne resistance. Number indicate numbers of resistance genes detected for each specific antibiotic subclass.

### CDIP genomic epidemiology using ONT vs. Illumina WGS

Similarly, a high level of diversity was expected in the chosen collection of clinical CDIP isolates with five different MLST sequence types and five clonal clusters based on Illumina assemblies (**Figure 8A**). Percentages of successfully detected cgMLST target genes in the assemblies were highest for Illumina (94.96% on average), but on par with the Nanopore approach SUPD50 (94.38 % on average) (**Figure 8B**).

**Figure 8.**
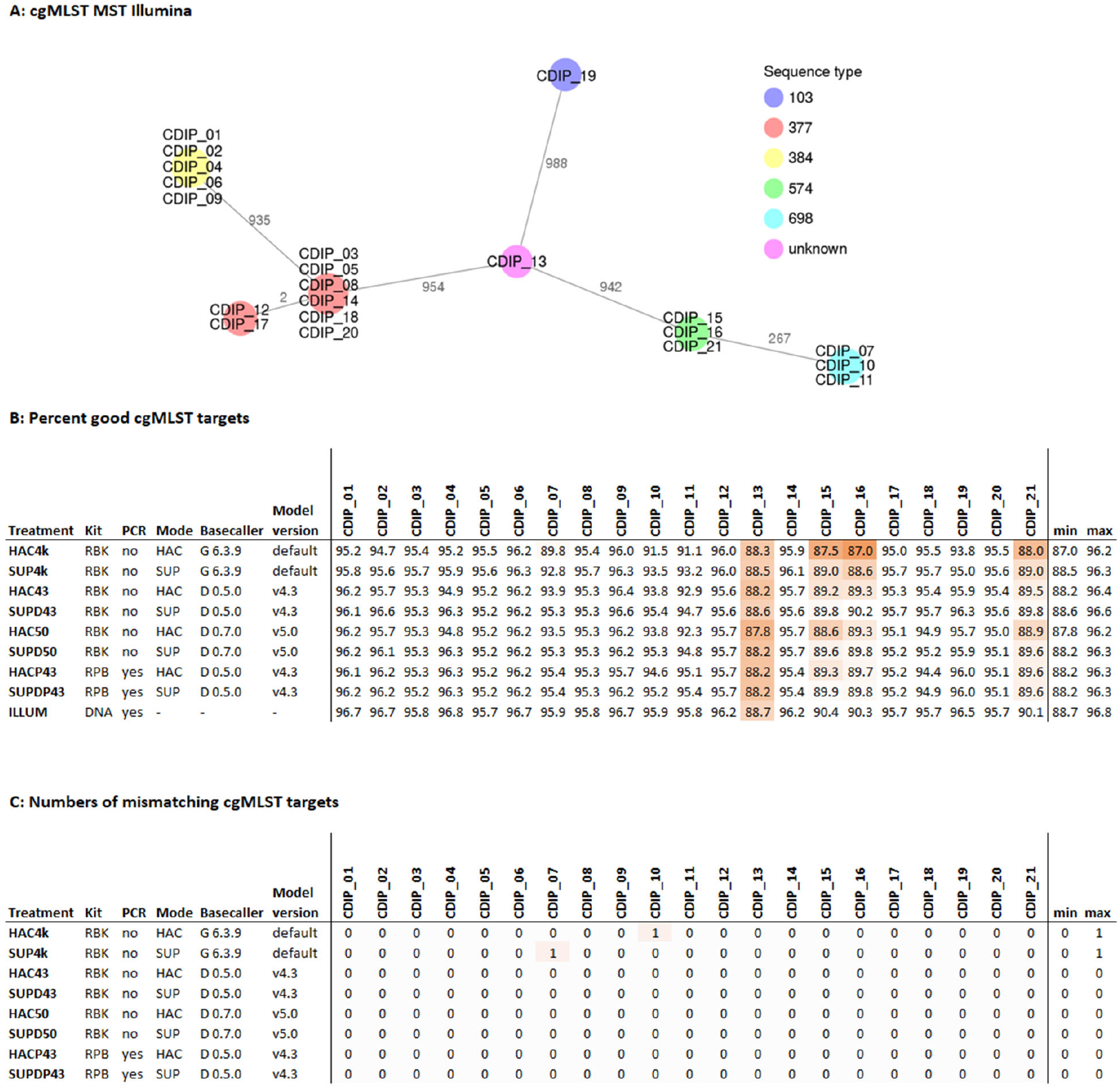
Comparison of cgMLST results from 21 clinical *Corynebacterium diphtheriae* isolates sequenced in parallel with Illumina and ONT. The cgMLST scheme contained 1,312 target genes. Treatment: Sequencing and data processing protocol used (Table S1). A) Minimum spanning tree of the 21 Illumina assemblies. B) Percentage the 1,312 cgMLST target genes that were detected (and passed the QC) in the different assemblies. C) Numbers of mismatching alleles between the ONT and the corresponding Illumina assemblies.

Masking of ambiguous base positions reduced the number of successfully detected cgMLST target genes by 1.1% and 1.5% in the treatments SUPD43.masked and HAC43.masked as compared to their unmasked counterparts SUPD43 and HAC43, respectively (**Figure S1**). Between Illumina and all tested Nanopore approaches, individual alleles were assigned in perfect agreement for MLST (**Figure S2C**), and with very high agreement for cgMLST, with a maximum of one differing allele per Nanopore approach (**Figure 8C**). The AMR predictions for CDIP showed complete concordance between Illumina and ONT assemblies at the class level (**Figure S3A)**. However, a single discrepancy was observed at the sub-class and gene levels for two isolates (CDIP_05 and CDIP_18). Specifically, the aminoglycoside phosphotransferase gene aph(3’)-Ia conferring kanamycin resistance was absent in the PCR-based ONT assemblies of both isolates (**Figures S3B, S3C**).

### Time and cost estimation

The ONT approaches used in this study are competitive with Illumina both in terms of hands-on time, overall turnaround time, and costs per sample at the tested throughput levels (**Table 1**). There is only a slight cost increase when switching from ONT RBK to RPB workflows, as the latter involves the use of long-range PCR in addition to the transposase library preparation as in the RBK protocol. Given the possibility to wash the ONT flow cells with a DNase treatment, and to stop the sequencing run earlier when enough reads have been obtained, a significant cost reduction can be obtained, as compared to the Illumina-based approach, which entails fixed costs and run durations.

**Table 1.**
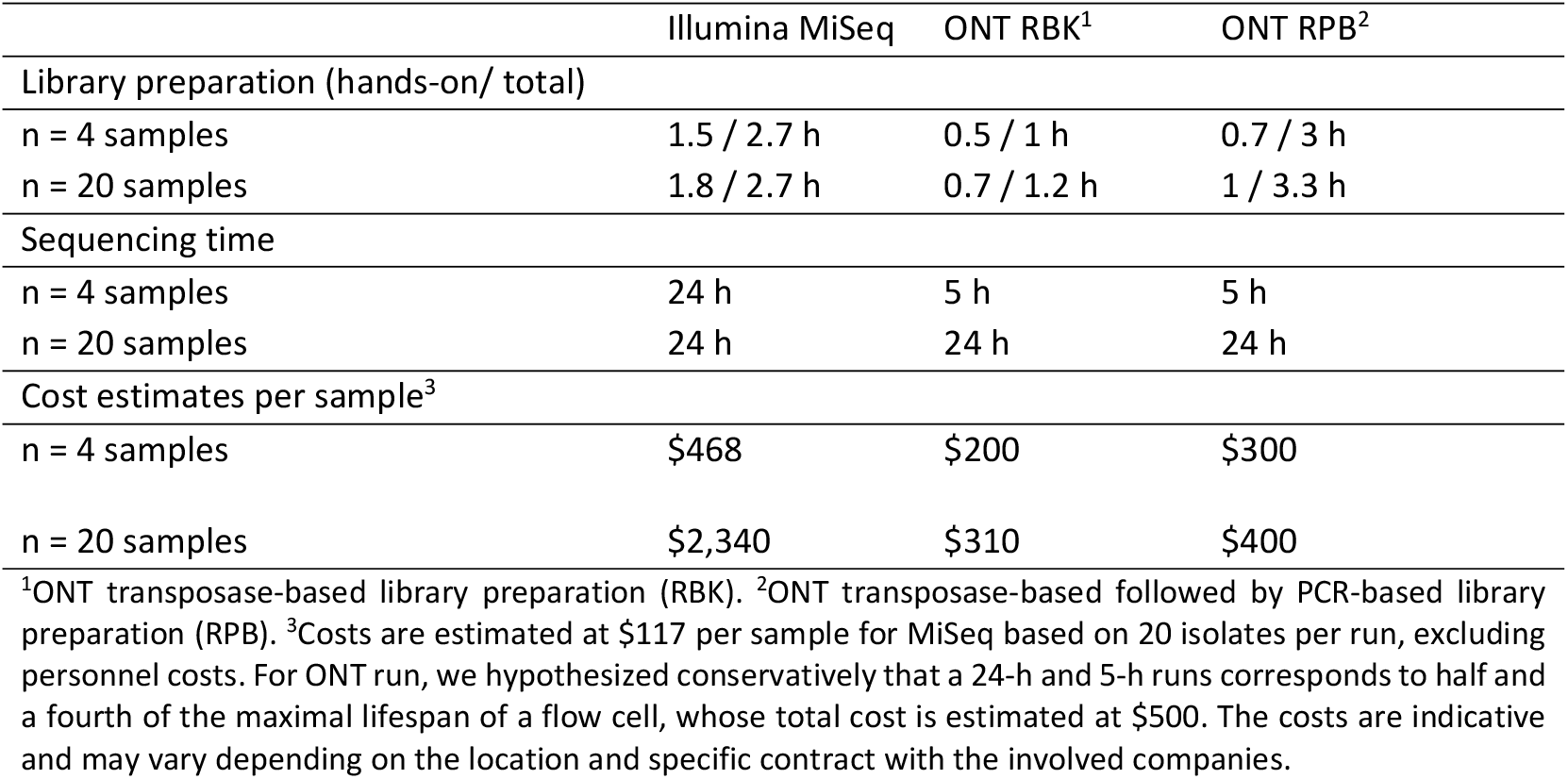
Turnaround time and cost estimates

## 4 Discussion

We assessed the recovery of cgMLST target genes, allele assignments, MLST and cgMLST classification, prediction of AMR genes and plasmid content using various Nanopore library preparation methods (PCR-based, PCR-free, and mixtures) and analysis pipelines. We evaluated a diverse, clinically relevant collection of outbreak isolates, originating from local outbreaks, which were classified as different MLST sequence types and clonal clusters based on Illumina WGS assemblies. Our main findings indicate that MLST classification was generally successful across Nanopore approaches, while cgMLST complex type (CT) classification showed more sensitivity to the different Nanopore approaches. We also explored the suitability of Nanopore assemblies for AMR and plasmid analysis, revealing generally concordant results with Illumina at the class level for AMR but some discrepancies at the subclass and gene levels, as well as in the genomic context of resistance genes. Finally, the study provided turnaround time and cost estimation, suggesting that the tested Nanopore approaches are competitive with those offered by short-read NGS approaches. With more than 550 resulting assemblies analyzed with MLST and cgMLST to determine clonal complexes and accuracy of the epidemiological conclusions, we highlighted how type of WGS data produced combined with the software running on Nanopore sequencers, as well as parameters selected by the operator, can have a major influence on the quality of the resulting sequencing reads and the epidemiological conclusions derived from the WGS data. Our results have practical consequences for genomic studies that aggregate sequencing data originating from different laboratories using different technologies or software versions.

While Nanopore demonstrated near-identical performance to Illumina for cgMLST target gene recovery and MLST typing, some discrepancies were observed in cgMLST allele assignments and AMR gene detection depending on the specific Nanopore approach used. Notably, PCR-based Nanopore library preparation generally outperformed PCR-free methods for allelic profiling in VRE. We concluded that optimized Nanopore sequencing and analysis pipelines offer a competitive alternative to Illumina for genomic surveillance of these pathogens in terms of accuracy, time, and cost. Complete concordance with Illumina results was observed for MLST in both tested species and for cgMLST in CDIP across all ONT kits and software evaluated. In contrast, the accuracy of cgMLST results for VRE varied based on the strain, library preparation kit, and analysis parameters, likely due to challenges in resolving base methylations. The latest software (Dorado 0.5.0) and basecalling model versions (≥ v4.3), combined with the PCR-based library preparation kit (RPB), reliably reproduced Illumina cgMLST results across all tested VRE strains.

In previous studies, ONT performance has been assessed for accurate genome reconstruction, however, most of these did not include the latest generation of flow cells, sequencing kits and software. For instance, Foster-Nyarko and colleagues (Foster-Nyarko et al., 2023) observed that MLST types and AMR determinants in *Klebsiella pneumoniae* could be reliably identified with R9.4.1 flow cells and V10 chemistry, but cluster detection (cgMLST, SNP) remained challenging with the approach. Using R9.4 flow cell and SQK-RBK001 kit, Greig et al. (Greig et al., 2019) found that 95% of the discrepant positions between Illumina and ONT sequencing data for two isolates of Shiga toxin–producing *Escherichia coli* (STEC) O157:H7 could be assigned to methylated positions in the genomes. Masking of the later combined with masking of prophage regions led to comparable results between Illumina and ONT. Using ONT R10.4 flow cells and V12 chemistry, a number of studies have further evaluated short and long-read WGS for accurate genome reconstruction (Linde et al., 2023; Sanderson et al., 2023; Sereika et al., 2022; Wagner et al., 2023): Sanderson et al. (Sanderson et al., 2023) concluded that the accuracy of the V12 chemistry was promising for high accuracy genome reconstruction, but that the throughput was insufficient for larger scale studies. They did not test cgMLST, but SNPs and indels for accuracy analysis. Linde et al. (Linde et al., 2023) benchmarked ONT V10 and V12 kits against Illumina by analyzing six strains of *B. anthracis, B. suis* and *F. tularensis* with cgMLST and cgSNP analyses, and concluded that high-resolution genotyping with V12 kits might be feasible for *F. tularensis* and *B. anthracis*, but not yet for *B. suis*. Sereika et al. (Sereika et al., 2022) analyzed seven bacterial species and concluded that R10.4 flow cells can generate near-finished bacterial genomes from pure cultures and metagenomes without the need for short-read or reference polishing. They defined the term “near finished” as a high-quality genome for which short-read polishing is not expected to significantly improve the consensus sequence. Yet, they did not use high-resolution typing schemes in their study. Finally, Wagner et al. (Wagner et al., 2023) benchmarked ONT V12 kits against Illumina short reads by analyzing 30 *Bordetella pertussis* isolates with cgMLST and concluded that ONT accuracy may be sufficient for clinical epidemiology. Overall, the previous generation of sequencing kits (R10.3 and R10.4 flow cells in combination with V12 chemistry) already showed significant improvements in accuracy, particularly in resolving homopolymer errors. Yet these were limited by their reduced throughput as compared to their predecessor (R9.4.1+ V10) and to the current ONT chemistry. In that regard, Lohde et al. evaluated the effectiveness of R10.4.1 flow cell and the latest V14 chemistry for outbreak detection with cgMLST, using isolates from a three-year-long *Klebsiella pneumoniae* outbreak (Lohde et al., 2024). They found that ONT sequencing resulted in considerable base errors, compared to Illumina data, leading to incorrect exclusions in outbreak tracing. They further identified the sequence motives around the putative methylation sites that lead to the inconsistencies, and suggested mitigating methylation related errors by ether using a PCR-based library preparation kit, or by masking ambiguous base positions.

Our study thus expands the knowledge on ONT performance for WGS-based bacterial typing by applying a similar comparative framework to two sets of well-characterized outbreak strains from the highly relevant species CDIP and VRE both with native and PCR-based library preparation kits. In contrast to previous studies, we put a strong focus on the influence of different sequencing and basecalling software on the accuracy of downstream analyses such as cgMLST and AMR gene contextual analyses. Thus, we tested multiple software versions released in 2023 and 2024, in combination with different parameters such as different versions of the GridION software, varying sampling rate of the signal acquisition (4khz and 5khz), basecaller software (Guppy and Dorado), basecaller versions and basecalling models. All sequencing runs were conducted with real time basecalling in high accuracy mode (HAC). The acquired signals were later re-basecalled in SUP mode, with standalone versions of the basecallers Guppy and Dorado, and different basecalling models. Our study thus reflects and integrates in its evaluation the rapid evolution of the technology over the past two years, including both wet laboratory and software changes that initially reduced the accuracy of the tested applications. We show the significant influence such update may have on ONT datasets, when processing native DNA libraries. Further strengths of our study are that we also provided estimates of both turnaround time and associated costs to offer a comprehensive perspective on implementation considerations.

Our study also identified several weaknesses and areas where ONT sequencing showed variability or discrepancies compared to Illumina. The number of mismatching alleles between Illumina and Nanopore assemblies varied strongly depending on the isolate, ONT software versions, basecalling models, and library preparation kits. PCR-free library preparation was found to be more sensitive to analysis parameters than PCR-based methods. Masking of ambiguous base positions with MPOA could lead to unassigned STs and a reduction in successfully detected cgMLST target genes. Discrepancies were observed in AMR predictions at the antibiotic subclass and gene levels for certain isolates. Furthermore, the predicted genomic context of resistance genes (chromosomal vs. plasmid-mediated) showed discrepancies between different Nanopore approaches and compared to Illumina in some cases. Despite its strengths, the PCR-based ONT kit exhibited certain drawbacks, such as smaller read length, ultimately leading to more fragmented chromosome assemblies. To mitigate this, we tested two hybrid strategies combining PCR-free and PCR-based Nanopore approaches and demonstrated promising results. These combined methods indeed retained the accuracy of the PCR-based approach while achieving assembly performance comparable to PCR-free methods. AMR prediction at the genome level was consistent across all tested sequencing methods, however resolving the genomic context of AMR genes remains a complex task (Djordjevic et al., 2024). Although ONT long-read sequencing may offer significant improvements in determining genomic context compared to short-read technologies, we did observe inconsistencies in the predicted genomic context of selected AMR genes, when comparing raw assemblies produced from PCR-based, PCR-free, and mixed Nanopore reads. As we chose a retrospective collection of outbreak-related isolates for in-depth comparative analyses, future work would need to provide a prospective evaluation, to assess the performance of the new technology in actual clinical or epidemiological settings, thus offering a more accurate reflection of real-world applicability. Also, we limited ourselves to local outbreaks, for which the source of material and reference short-read data were controlled. We may also ask how international collection and inter-laboratory comparison would perform in such a comparative framework.

The results of this study suggest that Nanopore WGS is a promising technology for genomic epidemiology of outbreak bacterial species, such as VRE and CDIP, demonstrating comparable performance to Illumina in key areas such as cgMLST target gene recovery and allele assignment. The strengths in chromosome assembly using PCR-free methods and the competitive time and cost further support this potential. However, the observed variability related to software and parameter specification and the discrepancies in AMR and plasmid analysis highlight the need for continued optimization of Nanopore sequencing protocols, basecalling algorithms, and analysis pipelines, to improve accuracy and reliability in genomic epidemiological applications. Future research could focus on refining PCR-free methods to reduce their sensitivity to analysis parameters, developing more robust methods for resolving ambiguous base positions, and improving the accuracy of AMR and plasmid prediction from Nanopore assemblies. As the technology continues to evolve, Nanopore sequencing has the potential to serve as a valuable alternative or complementary approach to short-read technologies for comprehensive genomic characterization in epidemiological investigations.

## Data Availability

http://www.ncbi.nlm.nih.gov/bioproject/1230056

https://github.com/RametteLab/NanoporeHybridKP

## References

Ahrenfeldt, J., Skaarup, C., Hasman, H., Pedersen, A.G., Aarestrup, F.M., Lund, O., 2017. Bacterial whole genome-based phylogeny: construction of a new benchmarking dataset and assessment of some existing methods. BMC Genomics 18, 19. 10.1186/s12864-016-3407-6

Arias, C.A., Murray, B.E., 2012. The rise of the Enterococcus: beyond vancomycin resistance. Nat Rev Microbiol 10, 266–278. 10.1038/nrmicro2761

Both, A., Kruse, F., Mirwald, N., Franke, G., Christner, M., Huang, J., Hansen, J.L., Kröger, N., Berneking, L., Lellek, H., Aepfelbacher, M., Rohde, H., 2022. Population dynamics in colonizing vancomycin-resistant Enterococcus faecium isolated from immunosuppressed patients. J Glob Antimicrob Resist 28, 267–273. 10.1016/j.jgar.2022.01.027

Cao, M.D., Ganesamoorthy, D., Elliott, A.G., Zhang, H., Cooper, M.A., Coin, L.J.M., 2016. Streaming algorithms for identification of pathogens and antibiotic resistance potential from real-time MinION(TM) sequencing. Gigascience 5, 32. 10.1186/s13742-016-0137-2

Danecek, P., Bonfield, J.K., Liddle, J., Marshall, J., Ohan, V., Pollard, M.O., Whitwham, A., Keane, T., McCarthy, S.A., Davies, R.M., Li, H., 2021. Twelve years of SAMtools and BCFtools. GigaScience 10, giab008. 10.1093/gigascience/giab008

de Been, M., Pinholt, M., Top, J., Bletz, S., Mellmann, A., van Schaik, W., Brouwer, E., Rogers, M., Kraat, Y., Bonten, M., Corander, J., Westh, H., Harmsen, D., Willems, R.J.L., 2015. Core Genome Multilocus Sequence Typing Scheme for High-Resolution Typing of Enterococcus faecium. J Clin Microbiol 53, 3788–3797. 10.1128/JCM.01946-15

De Coster, W., D’Hert, S., Schultz, D.T., Cruts, M., Van Broeckhoven, C., 2018. NanoPack: visualizing and processing long-read sequencing data. Bioinformatics 34, 2666–2669. 10.1093/bioinformatics/bty149

Deamer, D., Akeson, M., Branton, D., 2016. Three decades of nanopore sequencing. Nat Biotechnol 34, 518–524. 10.1038/nbt.3423

Delahaye, C., Nicolas, J., 2021. Sequencing DNA with nanopores: Troubles and biases. PLOS ONE 16, e0257521. 10.1371/journal.pone.0257521

Deurenberg, R.H., Bathoorn, E., Chlebowicz, M.A., Couto, N., Ferdous, M., García-Cobos, S., Kooistra-Smid, A.M.D., Raangs, E.C., Rosema, S., Veloo, A.C.M., Zhou, K., Friedrich, A.W., Rossen, J.W.A., 2017. Application of next generation sequencing in clinical microbiology and infection prevention. J Biotechnol 243, 16–24. 10.1016/j.jbiotec.2016.12.022

Didelot, X., Bowden, R., Wilson, D.J., Peto, T.E.A., Crook, D.W., 2012. Transforming clinical microbiology with bacterial genome sequencing. Nat Rev Genet 13, 601–612. 10.1038/nrg3226

Dilthey, A.T., Meyer, S.A., Kaasch, A.J., 2020. Ultraplexing: increasing the efficiency of long-read sequencing for hybrid assembly with k-mer-based multiplexing. Genome Biol 21, 68. 10.1186/s13059-020-01974-9

Djordjevic, S.P., Jarocki, V.M., Seemann, T., Cummins, M.L., Watt, A.E., Drigo, B., Wyrsch, E.R., Reid, C.J., Donner, E., Howden, B.P., 2024. Genomic surveillance for antimicrobial resistance — a One Health perspective. Nat Rev Genet 25, 142–157. 10.1038/s41576-023-00649-y

Feldgarden, M., Brover, V., Gonzalez-Escalona, N., Frye, J.G., Haendiges, J., Haft, D.H., Hoffmann, M., Pettengill, J.B., Prasad, A.B., Tillman, G.E., Tyson, G.H., Klimke, W., 2021. AMRFinderPlus and the Reference Gene Catalog facilitate examination of the genomic links among antimicrobial resistance, stress response, and virulence. Sci Rep 11, 12728. 10.1038/s41598-021-91456-0

Ferreira, F.A., Helmersen, K., Visnovska, T., Jørgensen, S.B., Aamot, H.V., 2021. Rapid nanopore-based DNA sequencing protocol of antibiotic-resistant bacteria for use in surveillance and outbreak investigation. Microb Genom 7, 000557. 10.1099/mgen.0.000557

Foster-Nyarko, E., Cottingham, H., Wick, R.R., Judd, L.M., Lam, M.M.C., Wyres, K.L., Stanton, T.D., Tsang, K.K., David, S., Aanensen, D.M., Brisse, S., Holt, K.E., 2023. Nanopore-only assemblies for genomic surveillance of the global priority drug-resistant pathogen, Klebsiella pneumoniae. Microb Genom 9. 10.1099/mgen.0.000936

Greig, D.R., Jenkins, C., Gharbia, S., Dallman, T.J., 2019. Comparison of single-nucleotide variants identified by Illumina and Oxford Nanopore technologies in the context of a potential outbreak of Shiga toxin–producing Escherichia coli. Gigascience 8, giz104. 10.1093/gigascience/giz104

Hall, M.B., Rabodoarivelo, M.S., Koch, A., Dippenaar, A., George, S., Grobbelaar, M., Warren, R., Walker, T.M., Cox, H., Gagneux, S., Crook, D., Peto, T., Rakotosamimanana, N., Grandjean Lapierre, S., Iqbal, Z., 2023. Evaluation of Nanopore sequencing for Mycobacterium tuberculosis drug susceptibility testing and outbreak investigation: a genomic analysis. The Lancet Microbe 4, e84– e92. 10.1016/S2666-5247(22)00301-9

Harris, S.R., 2018. SKA: Split Kmer Analysis Toolkit for Bacterial Genomic Epidemiology. 10.1101/453142

Higgs, C., Sherry, N.L., Seemann, T., Horan, K., Walpola, H., Kinsella, P., Bond, K., Williamson, D.A., Marshall, C., Kwong, J.C., Grayson, M.L., Stinear, T.P., Gorrie, C.L., Howden, B.P., 2022. Optimising genomic approaches for identifying vancomycin-resistant Enterococcus faecium transmission in healthcare settings. Nat Commun 13, 509. 10.1038/s41467-022-28156-4

Kofler, J., Ramette, A., Iseli, P., Stauber, L., Fichtner, J., Droz, S., Zihler Berner, A., Meier, A.B., Begert, M., Negri, S., Jachmann, A., Keller, P.M., Staehelin, C., Grützmacher, B., 2022. Ongoing toxin-positive diphtheria outbreaks in a federal asylum centre in Switzerland, analysis July to September 2022. Euro Surveill 27, 2200811. 10.2807/1560-7917.ES.2022.27.44.2200811

Kolmogorov, M., Yuan, J., Lin, Y., Pevzner, P.A., 2019. Assembly of long, error-prone reads using repeat graphs. Nat Biotechnol 37, 540–546. 10.1038/s41587-019-0072-8

Li, H., 2018. Minimap2: pairwise alignment for nucleotide sequences. Bioinformatics 34, 3094–3100. 10.1093/bioinformatics/bty191

Liao, Y.-C., Wu, H.-C., Liou, C.-H., Lauderdale, T.-L.Y., Huang, I.-W., Lai, J.-F., Chen, F.-J., 2022. Rapid and Routine Molecular Typing Using Multiplex Polymerase Chain Reaction and MinION Sequencer. Front Microbiol 13, 875347. 10.3389/fmicb.2022.875347

Linde, J., Brangsch, H., Hölzer, M., Thomas, C., Elschner, M.C., Melzer, F., Tomaso, H., 2023. Comparison of Illumina and Oxford Nanopore Technology for genome analysis of Francisella tularensis, Bacillus anthracis, and Brucella suis. BMC Genomics 24, 258. 10.1186/s12864-023-09343-z

Lohde, M., Wagner, G.E., Dabernig-Heinz, J., Viehweger, A., Braun, S.D., Monecke, S., Diezel, C., Stein, C., Marquet, M., Ehricht, R., Pletz, M.W., Brandt, C., 2024. Accurate bacterial outbreak tracing with Oxford Nanopore sequencing and reduction of methylation-induced errors. Genome Res. 34, 2039–2047. 10.1101/gr.278848.123

Magi, A., Semeraro, R., Mingrino, A., Giusti, B., D’Aurizio, R., 2018. Nanopore sequencing data analysis: state of the art, applications and challenges. Brief Bioinform 19, 1256–1272. 10.1093/bib/bbx062

Maiden, M.C., Bygraves, J.A., Feil, E., Morelli, G., Russell, J.E., Urwin, R., Zhang, Q., Zhou, J., Zurth, K., Caugant, D.A., Feavers, I.M., Achtman, M., Spratt, B.G., 1998. Multilocus sequence typing: a portable approach to the identification of clones within populations of pathogenic microorganisms. Proc Natl Acad Sci U S A 95, 3140–3145. 10.1073/pnas.95.6.3140

Maiden, M.C.J., Jansen van Rensburg, M.J., Bray, J.E., Earle, S.G., Ford, S.A., Jolley, K.A., McCarthy, N.D., 2013. MLST revisited: the gene-by-gene approach to bacterial genomics. Nat Rev Microbiol 11, 728–736. 10.1038/nrmicro3093

Miles, A., 2024. alimanfoo/pysamstats.

Nadon, C., Van Walle, I., Gerner-Smidt, P., Campos, J., Chinen, I., Concepcion-Acevedo, J., Gilpin, B., Smith, A.M., Man Kam, K., Perez, E., Trees, E., Kubota, K., Takkinen, J., Nielsen, E.M., Carleton, H., FWD-NEXT Expert Panel, 2017. PulseNet International: Vision for the implementation of whole genome sequencing (WGS) for global food-borne disease surveillance. Euro Surveill 22, 30544. 10.2807/1560-7917.ES.2017.22.23.30544

nanoporetech/medaka, 2024.

Oh, S., Nam, S.K., Chang, H.E., Park, K.U., 2022. Comparative Analysis of Short- and Long-Read Sequencing of Vancomycin-Resistant Enterococci for Application to Molecular Epidemiology. Front Cell Infect Microbiol 12, 857801. 10.3389/fcimb.2022.857801

Price, V., Ngwira, L.G., Lewis, J.M., Baker, K.S., Peacock, S.J., Jauneikaite, E., Feasey, N., 2023. A systematic review of economic evaluations of whole-genome sequencing for the surveillance of bacterial pathogens. Microbial Genomics 9, 000947. 10.1099/mgen.0.000947

Sanderson, N.D., Kapel, N., Rodger, G., Webster, H., Lipworth, S., Street, T.L., Peto, T., Crook, D., Stoesser, N., 2023. Comparison of R9.4.1/Kit10 and R10/Kit12 Oxford Nanopore flowcells and chemistries in bacterial genome reconstruction. Microb Genom 9, mgen000910. 10.1099/mgen.0.000910

Schadron, T., van den Beld, M., Mughini-Gras, L., Franz, E., 2024. Use of whole genome sequencing for surveillance and control of foodborne diseases: status quo and quo vadis. Front. Microbiol. 15. 10.3389/fmicb.2024.1460335

Schürch, A.C., Arredondo-Alonso, S., Willems, R.J.L., Goering, R.V., 2018. Whole genome sequencing options for bacterial strain typing and epidemiologic analysis based on single nucleotide polymorphism versus gene-by-gene-based approaches. Clin Microbiol Infect 24, 350–354. 10.1016/j.cmi.2017.12.016

Seemann, T., 2014. Prokka: rapid prokaryotic genome annotation. Bioinformatics 30, 2068–2069. 10.1093/bioinformatics/btu153

Sereika, M., Kirkegaard, R.H., Karst, S.M., Michaelsen, T.Y., Sørensen, E.A., Wollenberg, R.D., Albertsen, M., 2022. Oxford Nanopore R10.4 long-read sequencing enables the generation of near-finished bacterial genomes from pure cultures and metagenomes without short-read or reference polishing. Nat Methods 19, 823–826. 10.1038/s41592-022-01539-7

Simar, S.R., Hanson, B.M., Arias, C.A., 2021. Techniques in bacterial strain typing: past, present, and future. Current Opinion in Infectious Diseases 34, 339. 10.1097/QCO.0000000000000743

Tarumoto, N., Sakai, J., Sujino, K., Yamaguchi, T., Ohta, M., Yamagishi, J., Runtuwene, L.R., Murakami, T., Suzuki, Y., Maeda, T., Maesaki, S., 2017. Use of the Oxford Nanopore MinION sequencer for MLST genotyping of vancomycin-resistant enterococci. J Hosp Infect 96, 296–298. 10.1016/j.jhin.2017.02.020

Van Goethem, N., Descamps, T., Devleesschauwer, B., Roosens, N.H.C., Boon, N.A.M., Van Oyen, H., Robert, A., 2019. Status and potential of bacterial genomics for public health practice: a scoping review. Implementation Science 14, 79. 10.1186/s13012-019-0930-2

Wagner, G.E., Dabernig-Heinz, J., Lipp, M., Cabal, A., Simantzik, J., Kohl, M., Scheiber, M., Lichtenegger, S., Ehricht, R., Leitner, E., Ruppitsch, W., Steinmetz, I., 2023. Real-Time Nanopore Q20+ Sequencing Enables Extremely Fast and Accurate Core Genome MLST Typing and Democratizes Access to High-Resolution Bacterial Pathogen Surveillance. Journal of Clinical Microbiology 61, e01631–22. 10.1128/jcm.01631-22

Wu, X., Luo, H., Ge, C., Xu, F., Deng, X., Wiedmann, M., Baker, R.C., Stevenson, A.E., Zhang, G., Tang, S., 2022. Evaluation of multiplex nanopore sequencing for Salmonella serotype prediction and antimicrobial resistance gene and virulence gene detection. Front Microbiol 13, 1073057. 10.3389/fmicb.2022.1073057

Xian, Z., Li, S., Mann, D.A., Huang, Y., Xu, F., Wu, X., Tang, S., Zhang, G., Stevenson, A., Ge, C., Deng, X., 2022. Subtyping Evaluation of Salmonella Enteritidis Using Single Nucleotide Polymorphism and Core Genome Multilocus Sequence Typing with Nanopore Reads. Appl Environ Microbiol 88, e0078522. 10.1128/aem.00785-22

