## Supplementary Material for "Evaluation of Oxford Nanopore Technologies workflows for genomic epidemiology of outbreak‐associated bacterial isolates in the clinical setting"

### Supplementary Figures

**A: Percent good cgMLST targets**

| | VRE_16 | VRE_14 | VRE_12 | VRE_21 | VRE_02 | VRE_04 | VRE_05 | VRE_18 | VRE_07 | VRE_08 | VRE_11 | VRE_10 | VRE_17 | VRE_19 | VRE_20 | VRE_03 | VRE_09 | VRE_01 | VRE_15 | VRE_06 | cour | mean | std | min | 25% | 50% | 75% | max |
|---|---|---|---|---|---|---|---|---|---|---|---|---|---|---|---|---|---|---|---|---|---|---|---|---|---|---|---|---|
| HAC4k | 98.7 | 99.4 | 99.2 | 99.0 | 99.1 | 99.2 | 98.7 | 97.9 | 99.2 | 99.0 | 99.3 | 99.3 | 97.7 | 94.7 | 96.3 | - | - | 98.3 | 98.2 | 98.3 | 19 | 98.2 | 1.6 | 93.6 | 98.1 | 98.7 | 99.2 | 99.4 |
| SUP4k | 98.7 | 99.4 | 99.2 | 99.0 | 99.2 | 99.2 | 98.7 | 98.0 | 99.1 | 99.2 | 99.3 | 99.2 | 97.7 | 94.9 | 97.5 | - | - | 98.5 | 98.7 | 98.6 | 19 | 98.5 | 1.1 | 94.9 | 98.2 | 98.7 | 99.2 | 99.4 |
| HAC5k | 98.6 | 99.4 | 99.2 | 99.0 | 99.2 | 99.2 | 98.5 | 98.0 | 99.2 | 99.1 | 99.2 | 99.2 | 97.7 | 94.9 | 99.0 | 98.2 | 98.4 | 98.1 | 98.7 | 98.7 | 20 | 98.6 | 1.0 | 94.9 | 98.3 | 98.8 | 99.2 | 99.4 |
| SUPD5k | 98.7 | 99.3 | 99.2 | 98.9 | 99.2 | 99.2 | 98.6 | 98.0 | 99.1 | 99.2 | 99.2 | 99.2 | 97.5 | 94.9 | 99.0 | 98.2 | 98.4 | 98.2 | 98.5 | 98.9 | 20 | 98.6 | 1.0 | 94.9 | 98.3 | 98.9 | 99.2 | 99.3 |
| SUPD.mod | 98.7 | 99.3 | 99.2 | 98.9 | 99.2 | 99.2 | 98.6 | 97.9 | 99.1 | 99.2 | 99.2 | 99.3 | 97.7 | 94.8 | 99.0 | 98.2 | 98.3 | 98.2 | 98.5 | 98.8 | 20 | 98.6 | 1.0 | 94.8 | 98.3 | 98.8 | 99.2 | 99.3 |
| SUPR | 98.6 | 99.4 | 99.2 | 99.0 | 99.2 | 99.1 | 98.8 | 97.8 | 99.2 | 99.2 | 99.0 | 99.3 | 97.9 | 94.9 | 99.0 | 98.7 | 98.6 | 98.7 | 98.8 | 99.2 | 20 | 98.7 | 1.0 | 94.9 | 98.6 | 99.0 | 99.2 | 99.4 |
| HAC43 | 98.6 | 99.2 | 99.2 | 99.0 | 99.0 | 99.2 | 98.7 | 98.0 | 99.2 | 99.2 | 99.3 | 99.2 | 97.4 | 94.7 | 99.0 | 98.3 | 98.5 | 98.5 | 98.8 | 98.9 | 20 | 98.6 | 1.0 | 94.7 | 98.5 | 98.9 | 99.2 | 99.3 |
| SUPD43 | 98.7 | 99.3 | 99.2 | 98.9 | 99.2 | 99.2 | 98.8 | 97.9 | 99.0 | 99.2 | 99.1 | 99.3 | 97.9 | 94.9 | 99.0 | 98.5 | 98.7 | 98.8 | 99.0 | 99.2 | 20 | 98.7 | 1.0 | 94.9 | 98.7 | 99.0 | 99.2 | 99.3 |
| HAC50 | 98.6 | 99.2 | 99.2 | 99.0 | 99.2 | 99.2 | 98.7 | 97.9 | 99.2 | 99.0 | 99.2 | 99.2 | 97.7 | 94.8 | 98.9 | 98.6 | 98.5 | 98.4 | 98.6 | 98.8 | 20 | 98.6 | 1.0 | 94.8 | 98.6 | 98.8 | 99.2 | 99.2 |
| SUPD50 | 98.6 | 99.4 | 99.2 | 99.0 | 99.2 | 99.2 | 98.8 | 97.9 | 99.2 | 99.2 | 99.2 | 99.3 | 97.9 | 94.9 | 99.0 | 98.8 | 98.5 | 98.8 | 99.0 | 99.3 | 20 | 98.7 | 1.0 | 94.9 | 98.8 | 99.0 | 99.2 | 99.4 |
| HAC43.masked | 98.2 | 99.0 | 98.8 | 98.7 | 98.9 | 98.9 | 98.0 | 97.5 | 98.1 | 98.5 | 99.0 | 98.7 | 95.6 | 93.6 | 98.5 | 95.6 | 95.2 | 96.1 | 96.1 | 96.2 | 20 | 97.5 | 1.6 | 93.6 | 96.1 | 98.2 | 98.7 | 99.0 |
| SUPD43.masked | 98.7 | 99.3 | 99.1 | 99.0 | 99.2 | 99.0 | 98.0 | 97.7 | 98.2 | 98.5 | 98.7 | 98.2 | 96.9 | 93.8 | 98.9 | 96.8 | 96.6 | 96.8 | 97.4 | 97.3 | 20 | 97.9 | 1.3 | 93.8 | 97.2 | 98.2 | 98.9 | 99.3 |
| HACP43 | 98.6 | 99.4 | 99.1 | 98.9 | 99.1 | 99.2 | 98.7 | 97.8 | 99.2 | 99.2 | 99.2 | 99.2 | 97.8 | 94.8 | 99.0 | 98.8 | 98.8 | 98.8 | 99.0 | 99.2 | 21 | 98.7 | 1.0 | 94.8 | 98.8 | 99.0 | 99.2 | 99.4 |
| SUPDP43 | 98.7 | 99.4 | 99.2 | 99.0 | 99.1 | 99.2 | 98.8 | 97.9 | 99.2 | 99.2 | 99.2 | 99.3 | 97.9 | 94.9 | 99.0 | 98.9 | 98.9 | 98.9 | 99.0 | 99.4 | 21 | 98.8 | 1.0 | 94.9 | 98.9 | 99.0 | 99.2 | 99.4 |
| SUPD&P43 | 98.7 | 99.4 | 99.2 | 98.8 | 99.2 | 99.2 | 98.8 | 97.8 | 99.2 | 99.2 | 99.3 | 99.3 | 98.0 | 94.9 | 99.0 | 98.9 | 98.8 | 98.7 | 99.0 | 99.4 | 20 | 98.7 | 1.0 | 94.9 | 98.8 | 99.0 | 99.2 | 99.4 |
| HAC43CC | 98.6 | 99.2 | 99.2 | 99.0 | 99.0 | 99.2 | 98.7 | 98.0 | 99.2 | 99.2 | 99.3 | 99.2 | 97.5 | 94.7 | 99.0 | 98.4 | 98.6 | 98.5 | 98.8 | 99.0 | 20 | 98.6 | 1.0 | 94.7 | 98.6 | 99.0 | 99.2 | 99.3 |
| SUPD43CC | 98.7 | 99.3 | 99.2 | 98.9 | 99.2 | 99.2 | 98.8 | 97.9 | 99.0 | 99.2 | 99.1 | 99.3 | 97.9 | 94.9 | 99.0 | 98.5 | 98.7 | 98.8 | 99.0 | 99.2 | 20 | 98.7 | 1.0 | 94.9 | 98.7 | 99.0 | 99.2 | 99.3 |
| ILLUM | 98.7 | 99.4 | 99.2 | 99.0 | 99.2 | 99.2 | 98.8 | 98.3 | 99.2 | 99.2 | 99.3 | 99.3 | 98.0 | 94.9 | 99.0 | 98.9 | 98.9 | 98.9 | 99.0 | 99.4 | 21 | 98.8 | 1.0 | 94.9 | 98.9 | 99.0 | 99.2 | 99.4 |

**B: Number of alleles different from Illumina**

| Treatment | VRE_16 | VRE_14 | VRE_12 | VRE_21 | VRE_02 | VRE_04 | VRE_05 | VRE_18 | VRE_07 | VRE_08 | VRE_11 | VRE_10 | VRE_17 | VRE_19 | VRE_20 | VRE_03 | VRE_09 | VRE_01 | VRE_15 | VRE_06 | cour | mean | std | min | 25% | 50% | 75% | max |
|---|---|---|---|---|---|---|---|---|---|---|---|---|---|---|---|---|---|---|---|---|---|---|---|---|---|---|---|---|
| HAC4k | 0 | 0 | 0 | 0 | 1 | 0 | 0 | 1 | 4 | 4 | 5 | 4 | 6 | 6 | 19 | - | - | 27 | 20 | 32 | 19 | 7.2 | 9.8 | 0.0 | 0.0 | 4.0 | 7.0 | 32.0 |
| SUP4k | 0 | 0 | 0 | 0 | 0 | 0 | 2 | 1 | 2 | 2 | 1 | 4 | 6 | 11 | 16 | - | - | 44 | 39 | 39 | 19 | 9.0 | 14.7 | 0.0 | 0.0 | 2.0 | 8.5 | 44.0 |
| HAC5k | 0 | 0 | 0 | 0 | 0 | 1 | 1 | 1 | 6 | 5 | 9 | 7 | 4 | 6 | 0 | 11 | 16 | 8 | 15 | 14 | 20 | 5.2 | 5.5 | 0.0 | 0.0 | 4.5 | 8.3 | 16.0 |
| SUPD5k | 0 | 0 | 0 | 0 | 0 | 1 | 0 | 1 | 1 | 2 | 3 | 5 | 16 | 10 | 4 | 30 | 29 | 24 | 28 | 38 | 20 | 9.6 | 12.8 | 0.0 | 0.0 | 2.5 | 18.0 | 38.0 |
| SUPD.mod | 0 | 0 | 0 | 0 | 0 | 1 | 1 | 1 | 4 | 5 | 7 | 6 | 4 | 6 | 1 | 12 | 16 | 11 | 17 | 14 | 20 | 5.3 | 5.7 | 0.0 | 0.8 | 4.0 | 8.0 | 17.0 |
| SUPR | 0 | 0 | 0 | 0 | 0 | 0 | 1 | 1 | 0 | 0 | 0 | 0 | 5 | 3 | 0 | 10 | 13 | 14 | 13 | 16 | 20 | 3.8 | 5.8 | 0.0 | 0.0 | 0.0 | 6.3 | 16.0 |
| HAC43 | 0 | 0 | 0 | 0 | 0 | 0 | 0 | 1 | 1 | 0 | 0 | 0 | 11 | 9 | 3 | 22 | 30 | 19 | 19 | 27 | 20 | 7.1 | 10.4 | 0.0 | 0.0 | 0.5 | 13.0 | 30.0 |
| SUPD43 | 0 | 0 | 0 | 0 | 0 | 0 | 0 | 1 | 0 | 0 | 0 | 0 | 2 | 1 | 0 | 0 | 6 | 4 | 6 | 3 | 20 | 1.2 | 2.0 | 0.0 | 0.0 | 0.0 | 1.3 | 6.0 |
| HAC50 | 0 | 0 | 0 | 0 | 0 | 0 | 0 | 1 | 0 | 0 | 0 | 0 | 5 | 6 | 1 | 20 | 16 | 15 | 18 | 20 | 20 | 5.1 | 7.8 | 0.0 | 0.0 | 0.0 | 8.3 | 20.0 |
| SUPD50 | 0 | 0 | 0 | 0 | 0 | 0 | 1 | 2 | 0 | 1 | 0 | 0 | 0 | 0 | 0 | 0 | 4 | 1 | 0 | 1 | 20 | 0.5 | 1.0 | 0.0 | 0.0 | 0.0 | 1.0 | 4.0 |
| HAC43.masked | 0 | 0 | 0 | 0 | 0 | 0 | 0 | 1 | 0 | 0 | 0 | 0 | 0 | 1 | 0 | 2 | 4 | 3 | 2 | 3 | 20 | 0.8 | 1.3 | 0.0 | 0.0 | 0.0 | 1.3 | 4.0 |
| SUPD43.masked | 0 | 0 | 0 | 0 | 0 | 0 | 0 | 1 | 0 | 0 | 0 | 0 | 0 | 0 | 0 | 0 | 0 | 0 | 0 | 0 | 20 | 0.1 | 0.2 | 0.0 | 0.0 | 0.0 | 0.0 | 1.0 |
| HACP43 | 0 | 0 | 0 | 0 | 0 | 0 | 0 | 1 | 0 | 0 | 0 | 0 | 0 | 0 | 0 | 0 | 0 | 0 | 0 | 0 | 21 | 0.0 | 0.2 | 0.0 | 0.0 | 0.0 | 0.0 | 1.0 |
| SUPDP43 | 0 | 0 | 0 | 0 | 0 | 0 | 0 | 2 | 0 | 0 | 0 | 0 | 0 | 0 | 0 | 0 | 0 | 0 | 0 | 0 | 21 | 0.1 | 0.4 | 0.0 | 0.0 | 0.0 | 0.0 | 2.0 |
| SUPD&P43 | 0 | 0 | 0 | 0 | 0 | 0 | 0 | 2 | 0 | 0 | 0 | 0 | 0 | 0 | 0 | 0 | 0 | 0 | 0 | 0 | 20 | 0.1 | 0.4 | 0.0 | 0.0 | 0.0 | 0.0 | 2.0 |
| HAC43CC | 0 | 0 | 0 | 0 | 0 | 0 | 0 | 1 | 1 | 0 | 0 | 0 | 0 | 0 | 0 | 0 | 0 | 0 | 0 | 0 | 20 | 0.1 | 0.3 | 0.0 | 0.0 | 0.0 | 0.0 | 1.0 |
| SUPD43CC | 0 | 0 | 0 | 0 | 0 | 0 | 0 | 1 | 0 | 0 | 0 | 0 | 0 | 0 | 0 | 0 | 0 | 0 | 0 | 0 | 20 | 0.1 | 0.2 | 0.0 | 0.0 | 0.0 | 0.0 | 1.0 |

**C: MLST sequence type**

| Treatment | VRE_16 | VRE_14 | VRE_12 | VRE_21 | VRE_02 | VRE_04 | VRE_05 | VRE_18 | VRE_07 | VRE_08 | VRE_11 | VRE_10 | VRE_17 | VRE_19 | VRE_20 | VRE_03 | VRE_09 | VRE_01 | VRE_15 | VRE_06 |
|---|---|---|---|---|---|---|---|---|---|---|---|---|---|---|---|---|---|---|---|---|
| HAC4k | 117 | 117 | 117 | 117 | 78 | 117 | 796 | 80 | 117 | 117 | 117 | 117 | 133 | 296 | 117 | - | - | 796 | 796 | 117 |
| SUP4k | 117 | 117 | 117 | 117 | 78 | 117 | 796 | 80 | 117 | 117 | 117 | 117 | 133 | 296 | 117 | - | - | 796 | 796 | 117 |
| HAC5k | 117 | 117 | 117 | 117 | 78 | 117 | 796 | 80 | 117 | 117 | 117 | 117 | 133 | 296 | 117 | 796 | 796 | 796 | 796 | 117 |
| SUPD5k | 117 | 117 | 117 | 117 | 78 | 117 | 796 | 80 | 117 | 117 | 117 | 117 | 133 | 296 | 117 | 796 | 796 | 796 | 796 | 117 |
| SUPD.mod | 117 | 117 | 117 | 117 | 78 | 117 | 796 | 80 | 117 | 117 | 117 | 117 | 133 | 296 | 117 | 796 | 796 | 796 | 796 | 117 |
| SUPR | 117 | 117 | 117 | 117 | 78 | 117 | 796 | 80 | 117 | 117 | 117 | 117 | 133 | 296 | 117 | 796 | 796 | 796 | 796 | 117 |
| HAC43 | 117 | 117 | 117 | 117 | 78 | 117 | 796 | 80 | 117 | 117 | 117 | 117 | 133 | 296 | 117 | 796 | 796 | 796 | 796 | 117 |
| SUPD43 | 117 | 117 | 117 | 117 | 78 | 117 | 796 | 80 | 117 | 117 | 117 | 117 | 133 | 296 | 117 | 796 | 796 | 796 | 796 | 117 |
| HAC50 | 117 | 117 | 117 | 117 | 78 | 117 | 796 | 80 | 117 | 117 | 117 | 117 | 133 | 296 | 117 | 796 | 796 | 796 | 796 | 117 |
| SUPD50 | 117 | 117 | 117 | 117 | 78 | 117 | 796 | 80 | 117 | 117 | 117 | 117 | 133 | 296 | 117 | 796 | 796 | 796 | 796 | 117 |
| HAC43.masked | 117 | 117 | 117 | 117 | 78 | 117 | 796 | 80 | 117 | f | 117 | 117 | 133 | 296 | 117 | 796 | f | 796 | 796 | 117 |
| SUPD43.masked | 117 | 117 | 117 | 117 | 78 | 117 | 796 | 80 | 117 | 117 | 117 | 117 | 133 | 296 | 117 | 796 | 796 | 796 | 796 | 117 |
| HACP43 | 117 | 117 | 117 | 117 | 78 | 117 | 796 | 80 | 117 | 117 | 117 | 117 | 133 | 296 | 117 | 796 | 796 | 796 | 796 | 117 |
| SUPDP43 | 117 | 117 | 117 | 117 | 78 | 117 | 796 | 80 | 117 | 117 | 117 | 117 | 133 | 296 | 117 | 796 | 796 | 796 | 796 | 117 |
| SUPD&P43 | 117 | 117 | 117 | 117 | 78 | 117 | 796 | 80 | 117 | 117 | 117 | 117 | 133 | 296 | 117 | 796 | 796 | 796 | 796 | 117 |
| SUPD43C | 117 | 117 | 117 | - | 78 | 117 | 796 | 80 | 117 | 117 | 117 | 117 | 133 | 296 | 117 | 796 | 796 | 796 | 796 | 117 |
| SUPD43CC | 117 | 117 | 117 | - | 78 | 117 | 796 | 80 | 117 | 117 | 117 | 117 | 133 | 296 | 117 | 796 | 796 | 796 | 796 | 117 |
| ILLUM | 117 | 117 | 117 | 117 | 78 | 117 | 796 | 80 | 117 | 117 | 117 | 117 | 133 | 296 | 117 | 796 | 796 | 796 | 796 | 117 |

**D: cgMLST complex type**

| Treatment | VRE_16 | VRE_14 | VRE_12 | VRE_21 | VRE_02 | VRE_04 | VRE_05 | VRE_18 | VRE_07 | VRE_08 | VRE_11 | VRE_10 | VRE_17 | VRE_19 | VRE_20 | VRE_03 | VRE_09 | VRE_01 | VRE_15 | VRE_06 |
|---|---|---|---|---|---|---|---|---|---|---|---|---|---|---|---|---|---|---|---|---|
| HAC4k | 2094 | u | 1177 | 5602 | 1555 | 1177 | 1219 | 1552 | 4425 | 4424 | 1477 | u | 2888 | 426 | u | - | - | u | u | u |
| SUP4k | 2094 | u | 1177 | 5602 | 1555 | 1177 | 1219 | 1552 | 4425 | 4424 | 1477 | u | 2888 | 426 | u | - | - | | u | u |
| HAC5k | 2094 | u | 1177 | 5602 | 1555 | 1177 | 1219 | 1552 | 4425 | 4424 | 1477 | u | 2888 | 426 | 2505 | 2025 | u | 2025 | u | u |
| SUPD5k | 2094 | u | 1177 | 5602 | 1555 | 1177 | 1219 | 1552 | 4425 | 4424 | 1477 | u | 2888 | 426 | u | u | u | | u | u |
| SUPD.mod | 2094 | u | 1177 | 5602 | 1555 | 1177 | 1219 | 1552 | 4425 | 4424 | 1477 | u | 2888 | 426 | u | 2025 | | 2025 | u | u |
| SUPR | 2094 | u | 1177 | 5602 | 1555 | 1177 | 1219 | 1552 | 4425 | 4424 | 1477 | u | 2888 | 426 | 2505 | 2025 | 2025 | 2025 | u | u |
| HAC43 | 2094 | u | 1177 | 5602 | 1555 | 1177 | 1219 | 1552 | 4425 | 4424 | 1477 | u | 2888 | 426 | 7829 | | | | u | u |
| SUPD43 | 2094 | u | 1177 | 5602 | 1555 | 1177 | 1219 | 1552 | 4425 | 4424 | 1477 | u | 2888 | 426 | 2505 | 1219 | 2025 | 1219 | u | u |
| HAC50 | 2094 | u | 1177 | 5602 | 1555 | 1177 | 1219 | 1552 | 4425 | 4424 | 1477 | u | 2888 | 426 | 7829 | | | 2025 | u | u |
| SUPD50 | 2094 | u | 1177 | 5602 | 1555 | 1177 | 1219 | 2887 | 4425 | 4424 | 1477 | u | 564 | 426 | 2505 | 1219 | 1219 | 1219 | u | u |
| HAC43.masked | 2094 | u | 1177 | 5602 | 1555 | 1177 | 1219 | 1552 | 4425 | 4424 | 1477 | 24 | 564 | 426 | 2505 | 1219 | 1219 | 1219 | u | u |
| SUPD43.masked | 2094 | u | 1177 | 5602 | 1555 | 1177 | 1219 | 1552 | 4425 | 4424 | 1477 | 24 | 564 | 426 | 2505 | 1219 | 1219 | 1219 | u | u |
| HACP43 | 2094 | u | 1177 | 5602 | 1555 | 1177 | 1219 | 1552 | 4425 | 4424 | 1477 | u | 564 | 426 | 2505 | 1219 | 1219 | 1219 | u | u |
| SUPDP43 | 2094 | u | 1177 | 5602 | 1555 | 1177 | 1219 | 2887 | 4425 | 4424 | 1477 | u | 564 | 426 | 2505 | 1219 | 1219 | 1219 | u | u |
| SUPD&P43 | 2094 | u | 1177 | 5602 | 1555 | 1177 | 1219 | 2887 | 4425 | 4424 | 1477 | u | 564 | 426 | 2505 | 1219 | 1219 | 1219 | u | u |
| HAC43CC | 2094 | u | 1177 | 5602 | 1555 | 1177 | 1219 | 1552 | 4425 | 4424 | 1477 | u | 564 | 426 | 2505 | 1219 | 1219 | 1219 | u | u |
| SUPD43CC | 2094 | u | 1177 | 5602 | 1555 | 1177 | 1219 | 1552 | 4425 | 4424 | 1477 | u | 564 | 426 | 2505 | 1219 | 1219 | 1219 | u | u |
| ILLUM | 2094 | u | 1177 | 5602 | 1555 | 1177 | 1219 | 2887 | 4425 | 4424 | 1477 | u | 564 | 426 | 2505 | 1219 | 1219 | 1219 | u | u |

**Figure S1.** Comparison of cgMLST results for clinical VRE isolates sequenced with Illumina and Oxford Nanopore (ONT). A) Percentage the 1,423 cgMLST target genes that were detected (i.e. passed the QC) in the different assemblies. B) Numbers of mismatching alleles between the ONT and the corresponding Illumina assemblies. C) Detected MLST Sequence types (ST). The letters 'f' indicates failed analysis due to missing target genes. D) Complex types (cgMLST). The letter 'u' indicates unknown CT. Detailed information on the treatments is provided in Table S1.

**A: Percent good cgMLST targets**

| Treatment | CDIP_01 | CDIP_02 | CDIP_03 | CDIP_04 | CDIP_05 | CDIP_06 | CDIP_07 | CDIP_08 | CDIP_09 | CDIP_10 | CDIP_11 | CDIP_12 | CDIP_13 | CDIP_14 | CDIP_15 | CDIP_16 | CDIP_17 | CDIP_18 | CDIP_19 | CDIP_20 | CDIP_21 | count | mean | std | min | 25% | 50% | 75% | max |
|---|---|---|---|---|---|---|---|---|---|---|---|---|---|---|---|---|---|---|---|---|---|---|---|---|---|---|---|---|---|
| HAC43 | 96.2 | 95.7 | 95.3 | 94.9 | 95.2 | 96.2 | 93.9 | 95.3 | 96.4 | 93.8 | 92.9 | 95.6 | 88.2 | 95.7 | 89.2 | 89.3 | 95.3 | 95.4 | 95.9 | 95.4 | 89.5 | 21 | 94.0 | 2.6 | 88.2 | 93.8 | 95.3 | 95.7 | 96.4 |
| HAC43.masked | 95.6 | 94.9 | 94.9 | 94.2 | 94.8 | 95.6 | 93.6 | 95.0 | 95.1 | 93.3 | 90.6 | 95.3 | 86.1 | 95.4 | 88.0 | 88.4 | 94.9 | 94.7 | 93.1 | 94.4 | 88.7 | 21 | 93.2 | 2.9 | 86.1 | 93.1 | 94.7 | 95.0 | 95.6 |
| HAC4k | 95.2 | 94.7 | 95.43 | 95.2 | 95.5 | 96.19 | 89.8 | 95.43 | 96.0 | 91.5 | 91.1 | 95.96 | 88.3 | 95.88 | 87.5 | 87.0 | 95.0 | 95.5 | 93.8 | 95.5 | 88.0 | 21 | 93.3 | 3.3 | 87.0 | 91.1 | 95.2 | 95.5 | 96.2 |
| HACP43 | 96.1 | 96.2 | 95.27 | 96.3 | 95.2 | 96.19 | 95.4 | 95.27 | 95.7 | 94.6 | 95.1 | 95.66 | 88.2 | 95.35 | 89.3 | 89.7 | 95.2 | 94.4 | 96.0 | 95.1 | 89.6 | 21 | 94.3 | 2.6 | 88.2 | 94.6 | 95.3 | 95.7 | 96.3 |
| SUP4k | 95.8 | 95.6 | 95.73 | 95.9 | 95.58 | 96.27 | 92.8 | 95.66 | 96.3 | 93.5 | 93.2 | 95.96 | 88.5 | 96.11 | 89.0 | 88.6 | 95.7 | 95.7 | 95.0 | 95.6 | 89.0 | 21 | 94.1 | 2.8 | 88.5 | 93.2 | 95.6 | 95.8 | 96.3 |
| SUPD43 | 96.1 | 96.6 | 95.27 | 96.3 | 95.2 | 96.19 | 95.3 | 95.27 | 96.6 | 95.4 | 94.7 | 95.58 | 88.6 | 95.58 | 89.8 | 90.2 | 95.7 | 95.7 | 96.3 | 95.6 | 89.8 | 21 | 94.6 | 2.5 | 88.6 | 95.2 | 95.6 | 96.1 | 96.6 |
| SUPD43.masked | 95.5 | 95.6 | 94.9 | 95.2 | 94.8 | 95.7 | 94.2 | 95.0 | 95.9 | 94.1 | 93.1 | 95.3 | 87.0 | 95.4 | 88.4 | 88.6 | 95.0 | 94.9 | 91.4 | 94.8 | 88.7 | 21 | 93.5 | 2.8 | 87.0 | 93.1 | 94.9 | 95.3 | 95.9 |
| SUPDP43 | 96.2 | 96.2 | 95.2 | 96.3 | 95.2 | 96.2 | 95.4 | 95.3 | 96.2 | 95.2 | 95.4 | 95.7 | 88.2 | 95.4 | 89.9 | 89.8 | 95.2 | 94.9 | 96.0 | 95.1 | 89.6 | 21 | 94.4 | 2.6 | 88.2 | 95.1 | 95.3 | 96.0 | 96.3 |
| HAC50 | 96.2 | 95.7 | 95.3 | 94.8 | 95.2 | 96.2 | 93.5 | 95.3 | 96.2 | 93.8 | 92.3 | 95.7 | 87.8 | 95.7 | 88.6 | 89.3 | 95.1 | 94.9 | 95.7 | 95.0 | 88.9 | 21 | 93.8 | 2.8 | 87.8 | 93.5 | 95.1 | 95.7 | 96.2 |
| SUPD50 | 96.2 | 96.1 | 95.3 | 96.3 | 95.2 | 96.2 | 95.3 | 95.3 | 96.2 | 95.3 | 94.8 | 95.7 | 88.2 | 95.7 | 89.6 | 89.8 | 95.2 | 95.2 | 95.9 | 95.1 | 89.6 | 21 | 94.4 | 2.6 | 88.2 | 95.1 | 95.3 | 95.9 | 96.3 |
| ILLUM | 96.7 | 96.7 | 95.8 | 96.8 | 95.7 | 96.7 | 95.9 | 95.8 | 96.7 | 95.9 | 95.8 | 96.2 | 88.7 | 96.2 | 90.4 | 90.3 | 95.7 | 95.7 | 96.5 | 95.7 | 90.1 | 21 | 95.0 | 2.6 | 88.7 | 95.7 | 95.8 | 96.5 | 96.8 |

**B: Number of alleles different from Illumina**

| Treatment | CDIP_01 | CDIP_02 | CDIP_03 | CDIP_04 | CDIP_05 | CDIP_06 | CDIP_07 | CDIP_08 | CDIP_09 | CDIP_10 | CDIP_11 | CDIP_12 | CDIP_13 | CDIP_14 | CDIP_15 | CDIP_16 | CDIP_17 | CDIP_18 | CDIP_19 | CDIP_20 | CDIP_21 | count | mean | std | min | 0.25 | 0.5 | 0.75 | max |
|---|---|---|---|---|---|---|---|---|---|---|---|---|---|---|---|---|---|---|---|---|---|---|---|---|---|---|---|---|---|
| HAC43 | 0 | 0 | 0 | 0 | 0 | 0 | 0 | 0 | 0 | 0 | 0 | 0 | 0 | 0 | 0 | 0 | 0 | 0 | 0 | 0 | 0 | 21 | 0 | 0 | 0 | 0 | 0 | 0 | 0 |
| HAC43.masked | 0 | 0 | 0 | 0 | 0 | 0 | 0 | 0 | 0 | 0 | 0 | 0 | 0 | 0 | 0 | 0 | 0 | 0 | 0 | 0 | 0 | 21 | 0 | 0 | 0 | 0 | 0 | 0 | 0 |
| HAC4k | 0 | 0 | 0 | 0 | 0 | 0 | 0 | 0 | 0 | 1 | 0 | 0 | 0 | 0 | 0 | 0 | 0 | 0 | 0 | 0 | 0 | 21 | 0.05 | 0.2 | 0 | 0 | 0 | 0 | 1 |
| HACP43 | 0 | 0 | 0 | 0 | 0 | 0 | 0 | 0 | 0 | 0 | 0 | 0 | 0 | 0 | 0 | 0 | 0 | 0 | 0 | 0 | 0 | 21 | 0 | 0 | 0 | 0 | 0 | 0 | 0 |
| SUP4k | 0 | 0 | 0 | 0 | 0 | 0 | 1 | 0 | 0 | 0 | 0 | 0 | 0 | 0 | 0 | 0 | 0 | 0 | 0 | 0 | 0 | 21 | 0.05 | 0.2 | 0 | 0 | 0 | 0 | 1 |
| SUPD43 | 0 | 0 | 0 | 0 | 0 | 0 | 0 | 0 | 0 | 0 | 0 | 0 | 0 | 0 | 0 | 0 | 0 | 0 | 0 | 0 | 0 | 21 | 0 | 0 | 0 | 0 | 0 | 0 | 0 |
| SUPD43.masked | 0 | 0 | 0 | 0 | 0 | 0 | 0 | 0 | 0 | 0 | 0 | 0 | 0 | 0 | 0 | 0 | 0 | 0 | 0 | 0 | 0 | 21 | 0 | 0 | 0 | 0 | 0 | 0 | 0 |
| SUPDP43 | 0 | 0 | 0 | 0 | 0 | 0 | 0 | 0 | 0 | 0 | 0 | 0 | 0 | 0 | 0 | 0 | 0 | 0 | 0 | 0 | 0 | 21 | 0 | 0 | 0 | 0 | 0 | 0 | 0 |
| HAC50 | 0 | 0 | 0 | 0 | 0 | 0 | 0 | 0 | 0 | 0 | 0 | 0 | 0 | 0 | 0 | 0 | 0 | 0 | 0 | 0 | 0 | 21 | 0 | 0 | 0 | 0 | 0 | 0 | 0 |
| SUPD50 | 0 | 0 | 0 | 0 | 0 | 0 | 0 | 0 | 0 | 0 | 0 | 0 | 0 | 0 | 0 | 0 | 0 | 0 | 0 | 0 | 0 | 21 | 0 | 0 | 0 | 0 | 0 | 0 | 0 |

**C: MLST sequence type**

| Treatment | CDIP_01 | CDIP_02 | CDIP_03 | CDIP_04 | CDIP_05 | CDIP_06 | CDIP_07 | CDIP_08 | CDIP_09 | CDIP_10 | CDIP_11 | CDIP_12 | CDIP_13 | CDIP_14 | CDIP_15 | CDIP_16 | CDIP_17 | CDIP_18 | CDIP_19 | CDIP_20 | CDIP_21 |
|---|---|---|---|---|---|---|---|---|---|---|---|---|---|---|---|---|---|---|---|---|---|
| HAC43 | 384 | 384 | 377 | 384 | 377 | 384 | 698 | 377 | 384 | 698 | 698 | 377 | u | 377 | 574 | 574 | 377 | 377 | 103 | 377 | 574 |
| HAC43.masked | 384 | 384 | 377 | 384 | 377 | 384 | 698 | 377 | 384 | 698 | 698 | 377 | u | 377 | 574 | 574 | 377 | 377 | 103 | 377 | 574 |
| HAC4k | 384 | 384 | 377 | 384 | 377 | 384 | 698 | 377 | 384 | 698 | 698 | 377 | u | 377 | 574 | 574 | 377 | 377 | 103 | 377 | 574 |
| HACP43 | 384 | 384 | 377 | 384 | 377 | 384 | 698 | 377 | 384 | 698 | 698 | 377 | u | 377 | 574 | 574 | 377 | 377 | 103 | 377 | 574 |
| SUP4k | 384 | 384 | 377 | 384 | 377 | 384 | 698 | 377 | 384 | 698 | 698 | 377 | u | 377 | 574 | 574 | 377 | 377 | 103 | 377 | 574 |
| SUPD43 | 384 | 384 | 377 | 384 | 377 | 384 | 698 | 377 | 384 | 698 | 698 | 377 | u | 377 | 574 | 574 | 377 | 377 | 103 | 377 | 574 |
| SUPD43.masked | 384 | 384 | 377 | 384 | 377 | 384 | 698 | 377 | 384 | 698 | 698 | 377 | u | 377 | 574 | 574 | 377 | 377 | 103 | 377 | 574 |
| SUPDP43 | 384 | 384 | 377 | 384 | 377 | 384 | 698 | 377 | 384 | 698 | 698 | 377 | u | 377 | 574 | 574 | 377 | 377 | 103 | 377 | 574 |
| HAC50 | 384 | 384 | 377 | 384 | 377 | 384 | 698 | 377 | 384 | 698 | 698 | 377 | u | 377 | 574 | 574 | 377 | 377 | 103 | 377 | 574 |
| SUPD50 | 384 | 384 | 377 | 384 | 377 | 384 | 698 | 377 | 384 | 698 | 698 | 377 | u | 377 | 574 | 574 | 377 | 377 | 103 | 377 | 574 |
| ILLUM | 384 | 384 | 377 | 384 | 377 | 384 | 698 | 377 | 384 | 698 | 698 | 377 | u | 377 | 574 | 574 | 377 | 377 | 103 | 377 | 574 |

**Figure S2.** Comparison of cgMLST results from 21 clinical *Corynebacterium diphtheriae* isolates sequenced in parallel with Illumina and ONT. A) Percentage the 1,312 cgMLST target genes that were detected (i.e. passed the QC) in the different assemblies. B) Numbers of mismatching alleles between the ONT and the corresponding Illumina assemblies. C) Detected MLST Sequence types (ST). The letters 'u' indicates unknown CT. Detailed information on the treatments is provided in Table S1.

A)

| | CDIP_03 | CDIP_05 | CDIP_07 | CDIP_08 | CDIP_10 | CDIP_11 | CDIP_12 | CDIP_13 | CDIP_14 | CDIP_15 | CDIP_16 | CDIP_17 | CDIP_18 | CDIP_19 | CDIP_20 | CDIP_21 |
|---|---|---|---|---|---|---|---|---|---|---|---|---|---|---|---|---|
| AMINOGLYCOSIDE | 111 | 111 | 111 | 111 | 111 | 111 | 111 | 111 | 111 | 111 | 111 | 111 | 111 | | 111 | 111 |
| BETA-LACTAM | | | | | | | 111 | | 111 | | | | | | | |
| LINCOSAMIDE/MACROLIDE/STREPTOGRAMIN | | | | | | | 111 | | 111 | | | | | 111 | | |
| PHENICOL | 111 | 111 | 111 | 111 | 111 | 111 | 111 | 111 | 111 | 111 | 111 | 111 | 111 | 111 | 111 | 111 |
| QUATERNARY AMMONIUM | 111 | 111 | | 111 | | | 111 | | 111 | | | 111 | 111 | | 111 | |
| SULFONAMIDE | 111 | 111 | 111 | 111 | 111 | 111 | 111 | 111 | 111 | 111 | 111 | 111 | 111 | | 111 | 111 |
| TETRACYCLINE | 111 | 111 | | 111 | | | 111 | | 111 | | | 111 | 111 | 111 | 111 | |
| TRIMETHOPRIM | 111 | 111 | | 111 | | | 111 | | 111 | | | 111 | 111 | | 111 | |

B)

| | CDIP_03 | CDIP_05 | CDIP_07 | CDIP_08 | CDIP_10 | CDIP_11 | CDIP_12 | CDIP_13 | CDIP_14 | CDIP_15 | CDIP_16 | CDIP_17 | CDIP_18 | CDIP_19 | CDIP_20 | CDIP_21 |
|---|---|---|---|---|---|---|---|---|---|---|---|---|---|---|---|---|
| AZITHROMYCIN/CLINDAMYCIN/ERYTHROMYCIN/LINCOMYCIN/STREPTOGRAMIN B/TYLOSIN | | | | | | | 111 | | 111 | | | | | 111 | | |
| BETA-LACTAM | | | | | | | 111 | | 111 | | | | | | | |
| CHLORAMPHENICOL | 111 | 111 | 111 | 111 | 111 | 111 | 111 | 111 | 111 | 111 | 111 | 111 | 111 | 111 | 111 | 111 |
| GENTAMICIN/KANAMYCIN/TOBRAMYCIN | | | | | | | 111 | | 111 | | | | | | | |
| KANAMYCIN | 111 | 011 | 111 | 111 | 111 | 111 | | 111 | | 111 | 111 | 111 | 011 | | 111 | 111 |
| QUATERNARY AMMONIUM | 111 | 111 | | 111 | | | 111 | | 111 | | | 111 | 111 | | 111 | |
| STREPTOMYCIN | 111 | 111 | 111 | 111 | 111 | 111 | 111 | 111 | 111 | 111 | 111 | 111 | 111 | | 111 | 111 |
| SULFONAMIDE | 111 | 111 | 111 | 111 | 111 | 111 | 111 | 111 | 111 | 111 | 111 | 111 | 111 | | 111 | 111 |
| TETRACYCLINE | 111 | 111 | | 111 | | | 111 | | 111 | | | 111 | 111 | 111 | 111 | |
| TRIMETHOPRIM | 111 | 111 | | 111 | | | 111 | | 111 | | | 111 | 111 | | 111 | |

C)

| | CDIP_03 | CDIP_05 | CDIP_07 | CDIP_08 | CDIP_10 | CDIP_11 | CDIP_12 | CDIP_13 | CDIP_14 | CDIP_15 | CDIP_16 | CDIP_17 | CDIP_18 | CDIP_19 | CDIP_20 | CDIP_21 |
|---|---|---|---|---|---|---|---|---|---|---|---|---|---|---|---|---|
| aadA1 | | | | | | | 111 | | 111 | | | | | | | |
| aadA5 | 111 | 111 | | 111 | | | 111 | | 111 | | | 111 | 111 | | 111 | |
| ant(2'')-Ia | | | | | | | 111 | | 111 | | | | | | | |
| aph(3'')-Ib | 111 | 111 | 111 | 111 | 111 | 111 | | 111 | | 111 | 111 | 111 | 111 | | 111 | 111 |
| aph(3')-Ia | 111 | 011 | 111 | 111 | 111 | 111 | | 111 | | 111 | 111 | 111 | 011 | | 111 | 111 |
| aph(6)-Id | 111 | 111 | 111 | 111 | 111 | 111 | | 111 | | 111 | 111 | 111 | 111 | | 111 | 111 |
| blaOXA-2 | | | | | | | 111 | | 111 | | | | | | | |
| cml | 111 | 111 | | 111 | | | 111 | | 111 | | | 111 | 111 | | 111 | |
| cmx | 111 | 111 | 111 | 111 | 111 | 111 | | 111 | | 111 | 111 | 111 | 111 | 111 | 111 | 111 |
| dfrA1 | 111 | 111 | | 111 | | | 111 | | 111 | | | 111 | 111 | | 111 | |
| erm(X) | | | | | | | 111 | | 111 | | | | | 111 | | |
| qacEdelta1 | 111 | 111 | | 111 | | | 111 | | 111 | | | 111 | 111 | | 111 | |
| sul1 | 111 | 111 | 111 | 111 | 111 | 111 | 111 | 111 | 111 | 111 | 111 | 111 | 111 | | 111 | 111 |
| tet(33) | 111 | 111 | | 111 | | | 111 | | 111 | | | 111 | 111 | | 111 | |
| tet(O) | | | | | | | | | | | | | | 111 | | |
| tet(W) | | | | | | | 111 | | 111 | | | | | | | |

**Figure S3.** Comparison of the resistance analysis results of ONT and Illumina based assemblies. Tuples indicate presence (1) or absence (0) in the treatments SUBDP_43 (RPB), SUPD_43 (RBK), and ILLUM (Illumina). A: Antibiotic classes, B: Antibiotic subclasses, C: Resistance genes.

### Supplementary Tables

**Table S1**. Overview of experimental parameters evaluated in the study.

| Samples | Treatment name | Kit | PCR | GridION release | Basecaller | Mode | Model | Sampling rate | Flye parameters | Medaka model |
|---|---|---|---|---|---|---|---|---|---|---|
| VRE | HAC4k | SQK-RBK114.96 | no | 22.10.7 | Guppy 6.3.9 | HAC | default, real-time basecalling | 4khz | --nano-raw | r1041_e82_260bps_hac_g632 |
| VRE | SUP4k | SQK-RBK114.96 | no | 22.10.7 | Guppy 6.3.9 | SUP | Default | 4khz | --nano-hq --read-error 0.03 | r1041_e82_260bps_sup_g632 |
| VRE | HAC5k | SQK-RBK114.96 | no | 23.04.6 | Guppy 6.5.7 | HAC | default, real-time basecalling | 5khz | --nano-raw | r1041_e82_400bps_hac_v4.2.0 |
| VRE | SUPD5k | SQK-RBK114.96 | no | 23.04.6 | Dorado 0.3.3 | SUP | dna_r10.4.1_e8.2_400bps_sup@v4.2.0’ | 5khz | --nano-hq --read-error 0.03 | r1041_e82_400bps_sup_v4.2.0 |
| VRE | SUPD.mod | SQK-RBK114.96 | no | 23.04.6 | Dorado 0.3.3 | SUP modified bases | dna_r10.4.1_e8.2_400bps_sup@v4.2.0' + 5mC +6mA | 5khz | --nano-hq --read-error 0.03 | r1041_e82_400bps_sup_v4.2.0 |
| VRE | SUPR | SQK-RBK114.96 | no | 23.04.6 | Dorado 0.4.0 | SUP | res_dna_r10.4.1_e8.2_400bps_sup@2023-09-22_bacterial-methylation | 5khz | --nano-hq --read-error 0.03 | r1041_e82_400bps_sup_v4.2.0 |
| VRE | HAC43 | SQK-RBK114.96 | no | 23.04.6 | Dorado 0.5.0 | HAC | dna_r10.4.1_e8.2_400bps_sup@v4.3.0’ | 5khz | --nano-raw | r1041_e82_400bps_hac_v4.3.0 |
| VRE | SUPD43 | SQK-RBK114.96 | no | 23.04.6 | Dorado 0.5.0 | SUP | dna_r10.4.1_e8.2_400bps_sup@v4.3.0’ | 5khz | --nano-hq --read-error 0.03 | r1041_e82_400bps_sup_v4.3.0 |
| VRE | HACP43 | SQK-RPB114.24 | yes | 23.11.7 | Dorado 0.5.0 | HAC | dna_r10.4.1_e8.2_400bps_sup@v4.3.0’ | 5khz | --nano-raw | r1041_e82_400bps_hac_v4.3.0 |
| VRE | SUPDP43 | SQK-RPB114.24 | yes | 23.11.7 | Dorado 0.5.0 | SUP | dna_r10.4.1_e8.2_400bps_sup@v4.3.0’ | 5khz | --nano-hq --read-error 0.03 | r1041_e82_400bps_sup_v4.3.0 |
| VRE | SUPD&P43 | 70% SQK-RBK114.96, 30% SQK-RPB114.24 | Previously produced sequencing reads were pooled before assembly. See SUPD43 and SUPDP43 above. | | | | | | | |
| VRE | HAC43CC | Assembly: SQK-RBK114.96, Correction: SQK-RPB114.24 | Previously produced assemblies were corrected with previously produced read sets. See HAC43 and HAC43 above. | | | | | | | |
| VRE | SUPD43CC | Assembly: SQK-RBK114.96, Correction: SQK-RPB114.24 | Previously produced assemblies were corrected with previously produced read sets. See SUPD43 and SUPDP43 above. | | | | | | | |
| VRE | SUPD50 | SQK-RBK114.96 | no | 23.04.6 | Dorado 0.7.0 | SUP | dna_r10.4.1_e8.2_400bps_sup@v5.0.0 | 5khz | --nano-hq --read-error 0.03 | r1041_e82_400bps_sup_v5.0.0' |
| VRE | HAC50 | SQK-RBK114.96 | no | 23.04.6 | Dorado 0.7.0 | HAC | dna_r10.4.1_e8.2_400bps_hac@v5.0.0 | 5khz | --nano-raw | r1041_e82_400bps_hac_v5.0.0' |
| VRE | ILLUM | Illumina DNA | no | - | - | 2 x 150 | - | - | | |
| CDIP | HAC4k | SQK-RBK114.96 | no | 22.12.5 | Guppy 6.4.6-1 | HAC | default, real-time basecalling | 4khz | --nano-raw | r1041_e82_260bps_hac_g632 |
| CDIP | SUP4k | SQK-RBK114.96 | no | 22.12.5 | Guppy 6.4.6-1 | SUP | default | 4khz | --nano-hq --read-error 0.03 | r1041_e82_260bps_sup_g632 |
| CDIP | HAC43 | SQK-RBK114.96 | no | 23.07.12, 23.07.12 | Dorado 0.5.0 | HAC | dna_r10.4.1_e8.2_400bps_sup@v4.3.0’ | 5khz | --nano-raw | r1041_e82_400bps_hac_v4.3.0 |
| CDIP | SUPD43 | SQK-RBK114.96 | no | 23.07.12, 23.07.12 | Dorado 0.5.0 | SUP | dna_r10.4.1_e8.2_400bps_sup@v4.3.0’ | 5khz | --nano-hq --read-error 0.03 | r1041_e82_400bps_sup_v4.3.0 |
| CDIP | ILLUM | Illumina DNA | no | - | - | 2 x 150 | - | - | | |

**Abbreviations**: VRE: Vancomycin resistant *Enterococcus*; CDIP: *Corynebacterium diphtheriae*; Treatment name: A string representing a specific combination of analysis parameters; HAC: High accuracy basecalling mode; SUP: Super high accuracy basecalling mode.
